# Modeling and Global Sensitivity Analysis of Strategies to Mitigate Covid-19 Transmission on a Structured College Campus

**DOI:** 10.1101/2022.04.01.22273316

**Authors:** Lihong Zhao, Fabian Santiago, Erica M. Rutter, Shilpa Khatri, Suzanne Sindi

**Affiliations:** Department of Applied Mathematics, University of California, Merced, 5200 North Lake Rd., Merced, CA, 95343, USA; Health Sciences Research Institute, University of California, Merced, 5200 North Lake Rd., Merced, CA, 95343, USA

## Abstract

In response to the COVID-19 pandemic, many higher educational institutions moved their courses on-line in hopes of slowing disease spread. The advent of multiple highly-effective vaccines offers the promise of a return to “normal” in-person operations, but it is not clear if—or for how long—campuses should employ non-pharmaceutical interventions such as requiring masks or capping the size of in-person courses. In this study, we develop and fine-tune a model of COVID-19 spread to UC Merced’s student and faculty population. We perform a global sensitivity analysis to consider how both pharmaceutical and non-pharmaceutical interventions impact disease spread. Our work reveals that vaccines alone may not be sufficient to eradicate disease dynamics and that significant contact with an infectious surrounding community will maintain infections on-campus. Our work provides a foundation for higher-education planning allowing campuses to balance the benefits of in-person instruction with the ability to quarantine/isolate infectious individuals.

## 1 Introduction

In late 2019, a novel coronavirus, SARS-CoV-2, was identified as the cause of a cluster of pneumonia cases [1]. On March 11, 2020 the World Health Organization declared the 2019 novel coronavirus outbreak (COVID-19) a pandemic [2]. Shortly after, nearly every higher-education institute rapidly transitioned all classes to on-line instruction to “flatten the epidemic curve”. As of October 13, 2022 the cumulative number of confirmed COVID-19 cases exceeds 620 million [3]. Although the availability of multiple effective vaccines offers the likelihood of a return to normal life, the emergence of highly-infectious variants and the advent of booster shots means that the return to our pre-COVID-19 existence is not in our immediate future [4].

Starting in Fall 2020 in the United States, colleges and universities have attempted to employ strategies to manage COVID-19 with mixed results. Overall, there were substantial increases in the number of new COVID-19 cases after school re-opening [5]. Moreover, even though, by age, college students are less likely to experience severe complications from COVID-19, the same is not true for their surrounding communities. During the winter of 2020, large surges in COVID-19 cases from college students were followed by subsequent infections and deaths in the wider community [6]. In addition, many campuses delayed their in-person instruction in early 2022 due to emergence of the omicron variant [7]. While there is a strong desire for higher-educational institutions to maintain inperson instruction, it is clear that for the foreseeable future this will require an effective COVID-19 management policy.

Nationally, educational institutions need to evaluate how to most effectively plan activities during the academic year while ensuring they do not contribute to local outbreaks [8, 9, 10]. Although some campuses, such as the University of California (UC) and California State University systems, are mandating the COVID-19 vaccine for all students and employees, these mandates will not be required by all campuses or campus populations [11]. In places where the COVID-19 vaccination is not mandated, the population vaccination levels are likely to vary with local COVID-19 vaccine acceptance patterns [12].

Mathematical models have a proven track record of providing novel insights into the spread and control of epidemics. Dynamic epidemic models have been used to study COVID-19 at many scales [13, 14, 15, 16, 17, 18, 19]. Given the wide-spread campus closures due to COVID-19, models have been developed to study the spread of COVID-19 on college campuses to evaluate reopening strategies [20, 21, 22]. Many such models have considered only student populations, thereby reducing the population to an isolated “bubble” in which students do not interact with individuals outside the campus and faculty are not accounted for. In this study, we develop a model for COVID-19 dynamics in “bubble-like” institutions—such as universities, nursing homes and prisons—where individuals within those communities have complex structured interactions defined by their roles but, rather than a bubble, the boundaries between these environments are porous and certain types of individuals—professors, staff, guards—intermix freely within a larger surrounding community where COVID-19 is also spreading. We began the modeling work we present during the summer of 2020, as our campus and other colleges around the world worked to manage instruction. While our initial underlying framework was developed before the emergence of multiple variants, we have modified our model to include the possibility of reinfection for both vaccinated and unvaccinated subpopulations.

In this study, we develop a structured SEIR model of COVID-19 dynamics on a college campus and investigate the sensitivity of behavior to the vaccinated population on campus and other non-pharmaceutical interventions (NPIs) such as mask-use and social distancing. Our goal is to understand how vaccine hesitancy both within the campus population and the surrounding community will impact disease propagation and which interventions will be the most effective. More specifically, we individually model the various subpopulations at the university, including on-campus undergraduates, off-campus undergraduates, graduate students, and faculty/staff. We connect our campus to the surrounding community where behavior outside the university will impact COVID-19 dynamics within the university. We perform a global sensitivity analysis of model behavior—cumulative number of infections at the end of the semester and infection doubling time—and consider the first and total-order effect of epidemic parameters and social contact behavior.

In Section 2, we first develop our structured epidemic model, then describe the model outputs we will study as well as the variance based sensitivity analysis approach we employ. In Section 3, we discuss the campus data we use to parameterize our model. Although we use the campus network of UC Merced, we believe our results are representative of mid-size rural colleges. In Section 4, we present our results. We conclude in Section 5. We note that under all conditions, NPIs are still important to mitigating the spread of COVID-19 on campus and urge universities to continue to support their use.

## 2 Methods

### 2.1 Model Description

In this section, we describe the ordinary differential equation (ODE) compartment model that we have used to make most of the analysis for the university as presented to the administration during the summer and fall of 2020, see Fig. 1(a). This SEIR model is unique as we model the four subpopulations: (1) undergraduate students living off campus, *u*, (2) undergraduate students living on-campus and in dorms, *d*, (3) graduate students, *g*, and (4) faculty and staff, *f*. Individuals are designated by both population and COVID-19 status. Fig. 1(b) presents the phases of the diseases for one of the subpopulations, the undergraduate students living off-campus (*u*).

**Figure 1:**
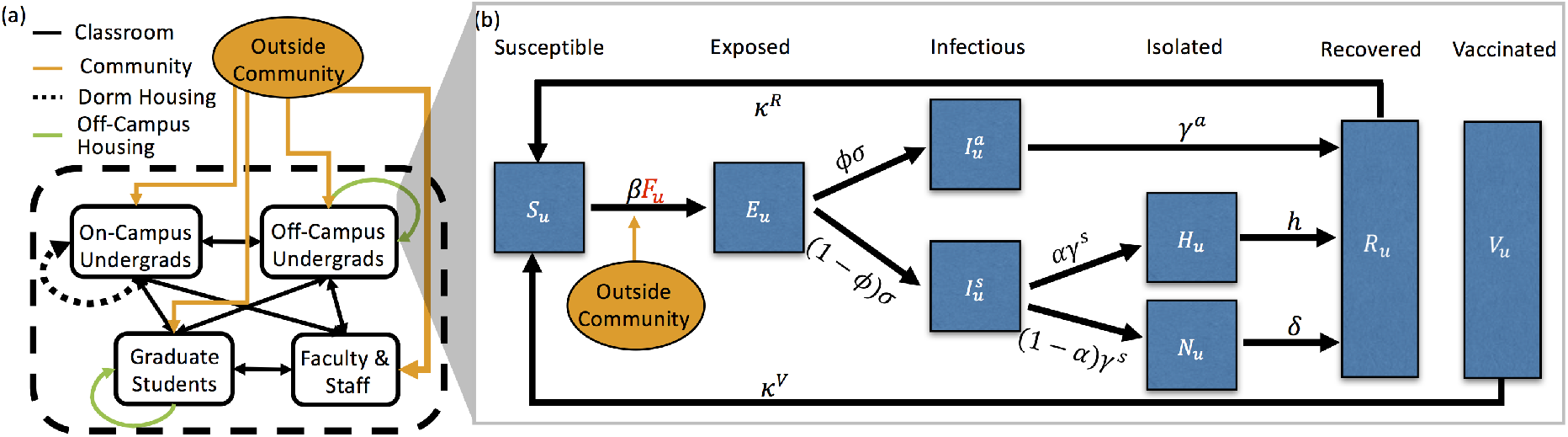
(a) Contacts between the 4 campus populations (on-campus undergraduates (*d*), off-campus undergraduates (*u*), graduate students (*g*), faculty/staff (*f*)) and the outside community. Contacts are separated into classroom (black), dormitory (black dotted), off-campus housing (green), and the outside community (orange). Thickness of arrows corresponds to the number of contact hours. (b) The stages of the COVID-19 infection, included in the ODE model, that an undergraduate living off-campus would progress through: susceptible (*S*), exposed (*E*), asymptomatically infectious (*I*^*a*^) or symptomatically infectious (*I*^*s*^), if symptomatically infectious individuals can choose to self-isolate (at home) (*H*) or not (*N*), and finally, both asymptomatically and symptomatically infectious individuals recover (*R*). Note that some percentage of the population is initially vaccinated (*V*) and, for simplicity, we assume no more individuals will get vaccinated once the semester begins. Both vaccinated individuals and recovered individuals can become susceptible after certain period of time

Each subpopulation in our model is divided into eight compartments related to different stages of infection and immune status: susceptible, *S*; exposed, *E*; asymptomatically infectious, *I*^*a*^; symptomatically infectious, *I*^*s*^; symptomatically infectious and in self-isolation (at home), *H*; symptomatically infectious but not in self-isolation, *N*; recovered, *R*; and vaccinated, *V*. A susceptible (*S*) individual may become exposed (*E*) to SARS-CoV-2 (the virus that causes COVID-19) by coming into contact with any infectious individuals (*I*) in any of the 4 subpopulations or the outside community (off campus). All exposed individuals become infectious, either asymptomatically (*I*^*a*^) with probability *φ* or symptomatically (*I*^*s*^) with probability 1 − *φ*, after spending an average of 1*/σ* days in exposed state. After an average of 1*/γ*^*s*^ days, symptomatic individuals may choose to self-isolate and/or report to health services for testing (*H*) with probability *α* or not (*N*) with probability 1 − *α*. Finally, as we do not model disease mortality, all sympotomatically infectious individuals eventually recover (*R*) — assuming recover rate for *H* and *N* individuals are *h* and *δ*, respectively. For asympotomatic individuals, we assume they recover after 1*/γ*^*a*^ days. For simplicity, we assume that both recovered individuals (*R*) and vaccinated individuals (*V*) are unable to contract the virus nor pass it to susceptible individuals for a certain period of time due to naturally acquired immunity (1*/κ*^*R*^) and vaccine-mediated immunity (1*/κ*^*V*^), respectively. We assume that the asymptomatically infectious individuals have lower probability in transmitting the disease than symptomatically infectious individuals, thus we employ the fraction of *β* of asymptomatically infectious individuals *ζ <* 1 where *β* denotes the transmission rate for symptomatically infectious individuals. Fig. 11 in Appendix A shows all the subpopulations and all the phases of the disease for each subpopulation.

For a subpopulation *i* ∈ {*u, d, g, f*}, the dynamics of each stage of the infection are governed by

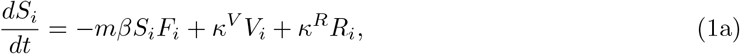

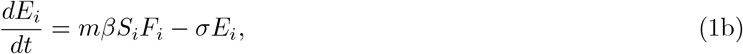

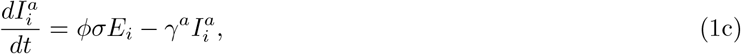

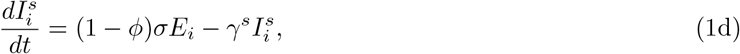

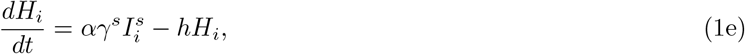

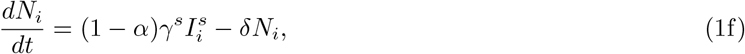

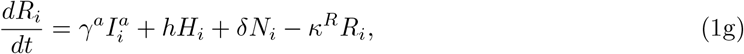

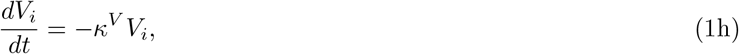

with initial conditions

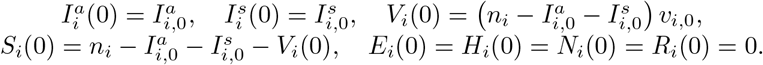

Here, *n*_*i*_ denotes the total number of individuals in subpopulation *i*, and *v*_*i*,0_ denotes the percentage of subpopulation *i* that has been vaccinated at time *t* = 0. Note that all the subpopulations are coupled through the force of infection, *F*_*i*_, where an individual in any subpopulation can be infected by any infectious individual in the campus population or through an infectious member of the community. The calculation involves the contact matrix ℂ,

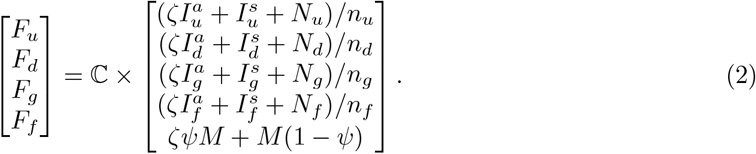

The contact matrix ℂ is a 4 × 5 matrix, with the indices 1, 2, 3, 4, and 5 correspond to off-campus undergraduate students, on-campus undergraduate students, graduate students, faculty/staff, and outside community, respectively. Taking off-campus undergraduate student as an example, we can obtain the explicit form of the force of infection *F*_*u*_ from Eq. (2):

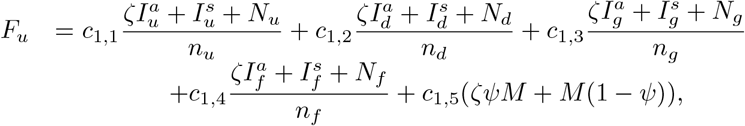

where *c*_*i,j*_ denotes the (*i, j*)-th entry of the contact matrix C. The first four terms signify the force of infection from off-campus undergraduate students, on-campus undergraduate students, graduate students, faculty/staff, respectively and are generated in the same fashion. For the first term, we can break it into the product between the number of contact-hours the off-campus undergraduate student has with other off-campus undergraduate students per day (*c*_1,1_), the probability of disease transmission per contact-hour (e.g. *ζ* if asymptomatic, 1 if symptomatic), and the proportion of contacts that are infectious (e.g., 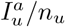 if asymptomatic, 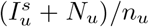 if symptomatic). The last term is generated by contact-hours between the average undergraduate student and the outside community per day (*c*_1,5_) multiplied by the static population of infectious individuals in the surrounding community (both asymptomatic and symptomatic), where *M* denotes the probability that an outside community individual is infectious and *ψ* denote the probability that an outside community individual with COVID-19 is asymptomatic.

The model has two types of parameters: (i) parameters related to COVID-19 epidemiology and (ii) parameters related to contact patterns between the 4 subpopulations and the outside community (contact matrix C is described in Table 2 and explained in Section 3). Details of the first type of parameters are given in Table 1. Details of contacts are given below in Section 3. The critical term in our model is the force of infection, defined in Eq. (2), which governs the spread of COVID-19 and is impacted by interventions such as mask-use, quarantine of infectious individuals, and changes in social connectivity, including class-size and housing caps through both types of parameters. We account for the effect of masks with our *mask efficiency* parameter which, when used in classrooms, reduced the transmission rate *β* by a factor (1 − *m*). In other words, *m* = 1 or 1 − *m* = 0 in Eqs. (1a) and (1b) corresponds to our baseline case when masks are not required or wearing a mask has no effect on the transmission rate *β*. We account for a quarantine period (1*/h*) and the probability that a symptomatic individual chose to self-isolate (*α*). Finally, changes in social connectivity, such as changes in class-sizes, are implemented by modifying the appropriate contact matrix.

**Table 1:** Description of parameters and their ranges used in the global sensitivity analysis. See Appendix A for details on parameterization.

| Symbol | Description | Unit | Range |
| --- | --- | --- | --- |
| $\beta$ | Transmission rate for symptomatic individuals per contact-hour (PCH) | PCH | $(5.74\text{--}24.86)\times 10^{-4}$ <sup>a</sup> |
| $1/\sigma$ | Expected time in exposed state | Day | 1–7.5 |
| $\phi$ | Probability that an exposed individual will become asymptomatic | — | 20%–80% |
| $\zeta$ | Fraction of $\beta$ for asymptomatic individuals | — | 50% |
| $1/\kappa^R$ | Average duration of naturally acquired immunity | Day | 90–270 |
| $1/\kappa^V$ | Average duration of vaccine-mediated immunity | Day | 180–270 |
| $M$ | Probability that outside (Merced) community individuals are (asymptomatically and symptomatically) infectious | — | 0%–1% |
| $\psi$ | Probability that outside (Merced) community individuals with COVID-19 are asymptomatic | — | 0.5 |
| $c$ | Community contact multiplier that multiplies contacts between university population and the community | — | 1–10 |
| $w$ | Weekend multiplier that scales the increase in social interaction during weekends | — | 1–10 |
| $p$ | Social percentage | — | 50%–150% |
| $1 - m$ | Reduction in $\beta$ with mask usage | — | 0%–60% |
| $\alpha$ | Probability that a symptomatic individual will self-isolate | — | 0%–100% |
| $1/\gamma^a$ | Average duration in asymptomatic infectious state | Day | 14 |
| $1/\gamma^s$ | Average duration in symptomatic state before deciding whether to isolate | Day | 2 |
| $1/h$ | Average duration of symptomatic individuals in self-isolation | Day | 12 |
| $1/\delta$ | Average duration in symptomatic state if not self-isolating | Day | 12 |
| $v_{u,0}, v_{d,0}$ | Percentage of $u$ or $d$ vaccinated at time $t = 0$ | — | 0%–80% |
| $v_{g,0}, v_{f,0}$ | Percentage of $g$ or $f$ vaccinated at time $t = 0$ | — | 0%–100% |
| $n_d$ | Number of on-campus undergraduates | People | 2885 |
| $n_u$ | Number of off-campus undergraduates | People | 5441 |
| $n_g$ | Number of graduates | People | 721 |
| $n_f$ | Number of faculty/staff | People | 424 |
| $I_{x,0}^y$ | Number of $y$ ($s = \text{symptomatic}$, $a = \text{asymptomatic}$) infectious individuals from population $x$ ($u = \text{off-campus undergrads}$, $d = \text{on-campus undergrads}$, $g = \text{graduate students}$, $f = \text{faculty \& staff}$) at time $t = 0$ | People | $(0\text{--}0.5\%) \times n_x$ |

<sup>a</sup> Our baseline value $\beta = 1.147 \times 10^{-3}$ was chosen so that a model of UC Merced, assuming classroom contacts based on the Fall 2019 schedule and housing contacts based on a proposed housing plan for Fall 2020 (before UC Merced was moved to fully-online), would propagate with an $R_0$ of 3 as defined by the next generation matrix approach [23].

**Table 2:** The contact matrix ℂ represents the total contact-hours per day within and between subpopulations, broken down by classes (ℂ^*c*^), living situations (ℂ^*l*^), outside community engagement (ℂ^*o*^), and unscheduled social interactions (ℂ^*s*^).

| Contacts | Off-Campus Under-graduates | On-Campus Under-graduates | Graduate Students | Faculty & Staff | Merced Community |
| --- | --- | --- | --- | --- | --- |
| Off-Campus Under-graduates | classroom + living + social | classroom + social | classroom | classroom | outside |
| On-Campus Under-graduates | classroom + social | classroom + living + social | classroom | classroom | outside |
| Graduate Students | classroom | classroom | classroom + living | classroom | outside |
| Faculty & Staff | classroom | classroom | classroom | faculty meetings | outside |

Because our goal is to investigate sensitivity of COVID-19 dynamics to NPIs in the context of a vaccinated campus population, we consider two simplifying assumptions allowing us to include vaccinated individuals in each of our campus subpopulations. First, our goal is to model COVID-19 in the context of a university environment. As such, we followed the requirements of the UC, which requires vaccination before the semester begins and did not consider an on-going vaccination program. Thus in our model we include all vaccinated individuals in the vaccinated compartment at time *t* = 0, and there is no inflow to the vaccinated compartment throughout the simulation. Second, we assume individuals would gain temporary immunity from vaccination which wanes over time. For simplicity, in our model, vaccinated individuals move to the susceptible compartment at the rate *κ*^*V*^. Similarly, we assume the immunity recovered individuals acquired from infection also wanes over time and thus in our model recovered individuals transition to the susceptible compartment at the rate *κ*^*R*^.

### 2.2 Infection Doubling Time

We are interested in analyzing the effect of intervention strategies and vaccination on the resulting dynamics of COVID-19 infections on our campus. In our work, the infection doubling time, Δ*T*, is the characteristic number of days for the cumulative number of COVID-19 infections to double, *C*(*t*_*i*+1_) = 2*C*(*t*_*i*_), where *t*_*i*+1_ = *t*_*i*_ + Δ*T* for *i ≥* 0 and *t*_0_ is a point in time at the beginning of the semester, see Fig. 2. This quantity is a characteristic of the disease dynamics during the early stages of disease spread (beginning of the semester), when the number of on-campus infections remains low, and the susceptible population remains large [24]. This characteristic captures the start of a semester when students that are allowed back to campus have been tested for COVID-19 (large susceptible population), and the probability that an infectious student returns to campus is low. Infection control measures aimed at “flattening the curve” means increasing the infection doubling time [25]. In this work, we use the infection doubling time as a measure of epidemic dynamics to asses which intervention strategies are associated with increased variance in the infection doubling time.

During this initial period of disease spread, at the beginning of a semester, we observe an exponential growth phase in the number of infections (Fig. 2). If the number of infections is growing at a rate *r*, the infection doubling time is given by Δ*T* = log(2)*/r* [26]. In this early stage of approximately exponential growth in the number of cumulative infection, *C*(*t*), where *C*′ (*t*) ≈ *rC*(*t*), we can estimate *r* by using cumulative infections at two points in time *C*(*t*_1_) and *C*(*t*_2_), where *r* = log (*C*(*t*_2_)*/C*(*t*_1_)) */*(*t*_2_ − *t*_1_) and *t*_1_ *< t*_2_. Then, the infection doubling time is

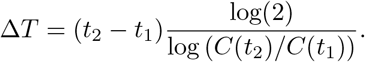

In this work, we use the average doubling time computed over consecutive days in the first month (four weeks) of the semester (see Fig. 2).

**Figure 2:**
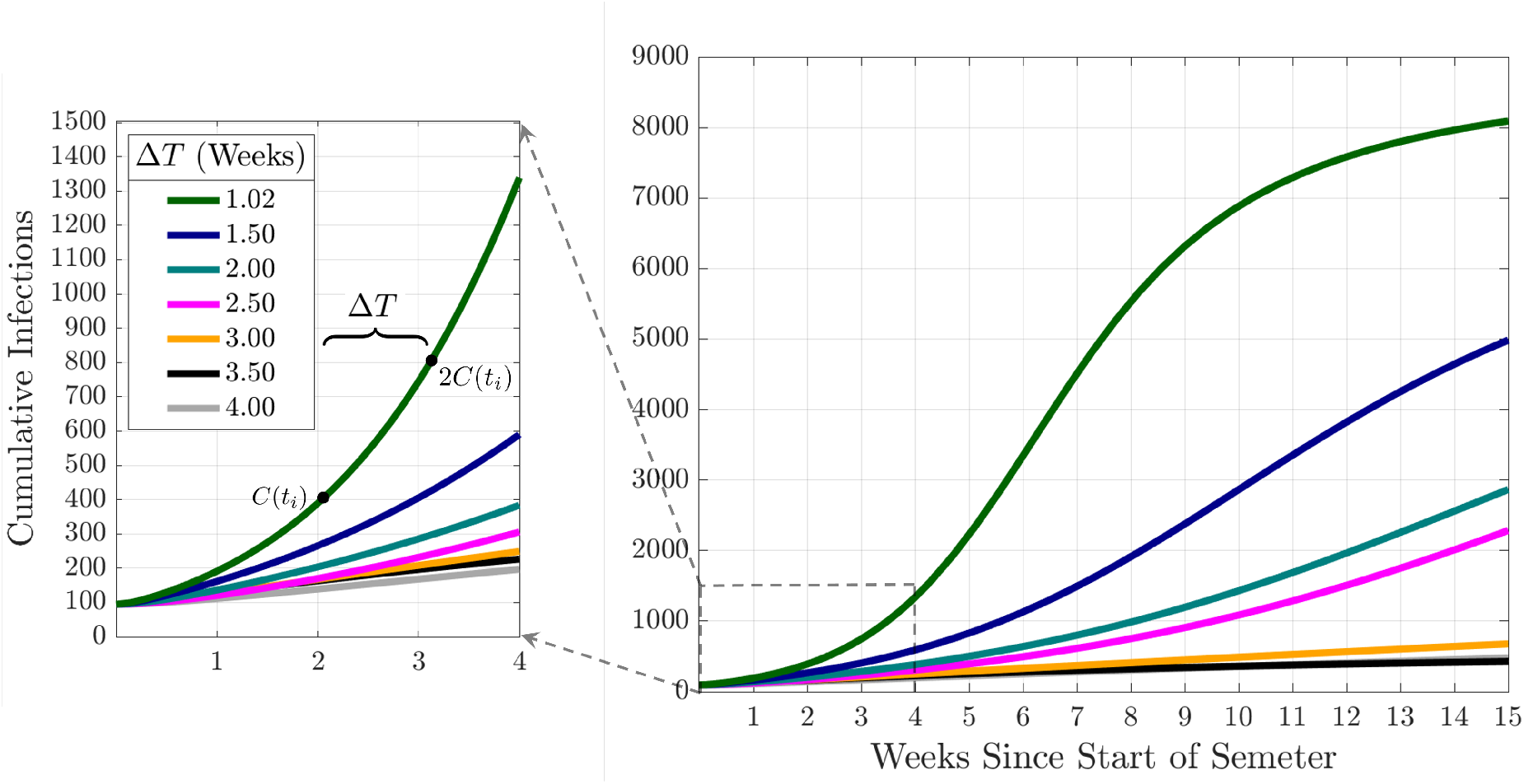
Cumulative Infections and Infection Doubling Times. The figure illustrates the doubling of the cumulative number of infections with respect to the infection doubling time (Δ*T*). Here, *C*(*t*_*i*_) is the cumulative number of infections at a time *t*_*i*_ since the beginning of the semester

### 2.3 Global Sensitivity Analysis

In this work we are interested in understanding how particular COVID-19 SEIR epidemic model factors ***θ*** = (*θ*_1_, *θ*_2_, …, *θ*_*k*_), Table 1, affect a model response *Y* (the cumulative number of infections, *C*(*t*), and the infection doubling time, Δ*T*). We perform a variance based global sensitivity analysis on these epidemic dynamics with the Sobol method [27, 28, 29], an approach to decompose the response variance by single and combined factor interactions

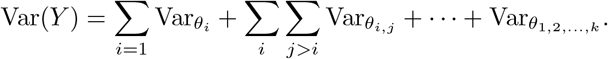

Under this decomposition, the proportion of variance from a single model factor, the *first-order* index, can be written as

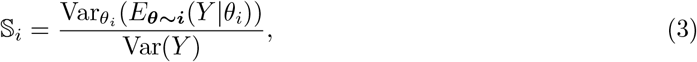

where *E* is expectation, Var is the variance, *θ*_*i*_ is the *i*^th^ model factor, and ***θ***_**∼*i***_ indicates varying all factors except *θ*_*i*_. In our work, a second quantity of interest is the *total-order* index, or total-effect index [30], which in addition to the first-order index information, accounts for the additional contribution of a factor to the model variance from interaction effects with other model factors

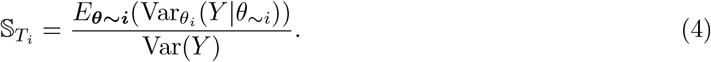

It follows that 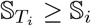, and that a total-order index value of zero indicates that the model factor is non-influential. In our analysis we estimate the first-order and total-order indices through numerical model solutions generated by sampling from the input factor parameter space following [27].

To estimate the first-order index, 𝕊_*i*_ (Eq. (3)), we use the estimator

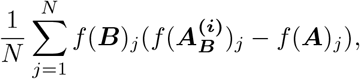

for a particular model factor *i*, first presented in [27]. Given a model response *Y* = *f* (·), where ***A*** and ***B*** are *N* × *k* matrices, in which each row is a sampling of the *k* model factors (*N* samples per matrix). Matrices ***A*** and ***B*** are generated using the same model factor sampling method and are thus interchangeable but serve for bookkeeping when forming the *k* matrices denoted by 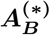. The matrix 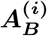 is an *N* × *k* matrix where column *i* comes from matrix ***B*** and all other *k* − 1 columns come from matrix ***A***.

The total-order index for a particular model factor *i*, 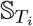 (Eq. (4)), is estimated with the estimator

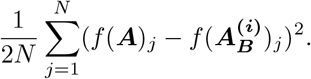

This estimator was first presented in [31]. For a more technical and detailed explanations of these estimators, one may consult [27].

To adequately sample the multidimensional input factor space, when forming the matrices ***A*** and ***B***, we use Latin hypercube sampling implemented in MATLAB using the Statistics and Machine Learning Toolbox [32]. To compute the first-order and total-order indices, we compute *N* (*k* + 2) = 31, 500 model solutions (*N* (*k* + 1) model solutions are suggested in [29] to estimate Sobol indices) with *N* = 1, 500 simulations for each of the *k* = 19 model factors. Each model factor is sampled from a uniform distribution over the range provided in Table 1. All confidence intervals (CIs) were formed by resampling the *N* (*k* + 2) simulations 2,000 times, with replacement, to compute the Sobol indices.

## 3 Data and Contacts

While the results we present here are specific to UC Merced, we note that any interested bubble-like communities could calculate their contacts as we outline below and use the model to analyze measures intended to mitigate COVID-19 transmission. For example, our model can be applied to skilled nursing facilities (where in-patients would be equivalent to ‘on-campus students’, out-patients would represent ‘off campus students’, doctors might correspond to ‘faculty/staff’, and therapists/nurses might translate to ‘graduate students’ and ‘classes’ in this case would be face-to-face treatments). UC Merced is a public land-grant university set in a rural community. In these calculations, there were 8,326 undergraduate students, of whom 2,885 live on-campus and 5,441 live off-campus. There were 721 graduate students and 424 faculty and staff. Lecturers and postdocs are considered part of the faculty/staff population. We expect our results would hold for similarly-sized campuses with similar class structures. Although we are able to get exact data from the registrar, we provide a template that can be used to estimate the contacts between different subpopulations.

For our model to be accurate, it is necessary to be able to estimate the number of contacts each of the subpopulations (off-campus undergraduates, on-campus undergraduates, graduate students, and faculty/staff) has with one another. We assume that most of these contacts come from in-class instruction, living/dorm situations, contact with the outside community, and unscheduled social interactions. We explain each of these factors separately and denote ℂ^*c*^ for classroom contacts, ℂ^*l*^ for living/dorm contacts, ℂ^*o*^ for contact with the outside community, and ℂ^*s*^ for unscheduled social interactions. The sum of these four matrices provides the total contact matrix ℂ. We use 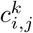 to denote the (*i, j*)-th entry of the submatrix ℂ^*k*^ with *k* ∈ {*c, l, o, s*}. For simplicity this information is displayed as a contact matrix in Table 2. We do not dynamically model the outside community and so the contact matrix is of size 4 × 5, since the interactions of campus population do not influence the outside community. The units of our contact matrix are contact-hours per day.

In the following subsections, we will discuss how the contact matrix is filled using in-class instruction, living situation, outside community interaction, and unscheduled social interaction information. Each of these sections corresponds to a different component of the contact matrix, as described in Table 2.

### 3.1 In-Class Instruction ℂ^*c*^

The majority of interactions in our model are derived from classroom instruction. We assume that lectures are comprised of 3 hours per week, while discussion sections meet for one hour per week. When a class is listed as a lab, we assume that it meets for 2.5 hours per week. Further, we assume that faculty and staff only interact with each other during faculty meetings. In particular, we assume a faculty meeting occurs once every other week for an hour. The ‘average’ department size was calculated using information from UC Merced School of Natural Sciences, and was roughly 17.5 faculty per department.

Especially in large classrooms, counting every other individual as a contact just as likely to spread COVID-19 is not realistic. COVID-19 is spread through droplets (e.g., sneezing, coughing) for contacts within 6 feet, but it can also be spread through aerosols which remain suspended in the air and viable for much longer [33, 34, 35]. As such, our formulation for the spread of COVID-19 in a classroom distinguishes between ‘close’ transmission (through droplets) and ‘far’ transmission (through aerosols). First, we assume that 8 contacts (the 8 closest people surrounding you) have a ‘normal’ chance of infecting you. Then, beyond your 8 neighbors, you have a reduced fraction (25%) of the ‘normal’ chance. (We also ran results with a 10% chance of infection, see Supplementary File 1, but the sensitivity results were qualitatively similar to the 25% chance.) As an example, assume an off-campus undergraduate student is in a 3-hour per week lecture course with 40 other students, 10 of whom live on-campus and 30 of whom live off-campus, then their ‘contact-hours’ from this one class with other off-campus undergraduate students would be

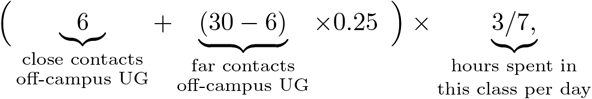

and with on-campus undergraduate students would be

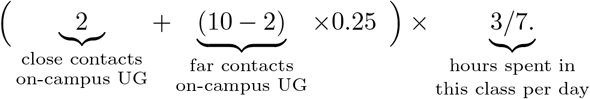

Note we assume that of the 8 contacts closest to the student, a proportional amount come from each sub-population. We compute these values for each of their courses and sum them together to get the total number for that student. We average over all the individuals in a subpopulation to obtain the corresponding classroom contact matrix entries.

We display the relevant information for calculating classroom contacts in Table 3. Classroom contacts only influence the campus subpopulations (not the outside community) and thus the fifth column (not displayed in Table 3) consists of all zeros. We note that in Table 3, the classes graduate students teach as graduate assistants may be labs or discussion sections, the classes faculty & staff teach may be lectures, labs, or discussion sections. For our baseline contact matrices and more details, see Appendix D.

**Table 3:** A schematic for calculating the contact due to teaching and class interactions at a university. Since classes only occur for the university community, this represents a 4 × 4 submatrix of Table 2. In other words, let ℂ^*cs*^ denote the matrix calculated from the schematic shown here, then the full classroom contact matrix would be 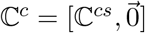 where 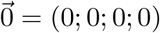.

| Classroom Contacts | Off-Campus Undergraduates | On-Campus Undergraduates | Graduate Students | Faculty & Staff |
| --- | --- | --- | --- | --- |
| Off-Campus Undergraduates | Off-campus students<br>per class ×<br># of classes taken ×<br>meeting hours per class | On-campus undergraduates<br>per class ×<br># of classes taken ×<br>meeting hours per class | # of classes taken ×<br>meeting hours per class | # of classes ×<br>meeting hours<br>per class |
| On-Campus Undergraduates | Off-campus students<br>per class ×<br># of classes taken ×<br>meeting hours per class | On-campus undergraduates<br>per class ×<br># of classes taken ×<br>meeting hours per class | # of classes taken ×<br>meeting hours per class | # of classes ×<br>meeting hours<br>per class |
| Graduate Students | Off-campus students<br>per class ×<br># of classes taught ×<br>meeting hours per class | On-campus undergraduates<br>per class ×<br># of classes taught ×<br>meeting hours per class | Graduates per class ×<br># of classes taken ×<br>meeting hours per class | # of classes ×<br>meeting hours<br>per meeting |
| Faculty & Staff | Off-campus students<br>per class ×<br># of classes taught ×<br>meeting hours per class | On-campus undergraduates<br>per class ×<br># of classes taught ×<br>meeting hours per class | Graduates per class ×<br># of classes taught ×<br>meeting hours per class | Size of<br>department ×<br>meeting hours<br>per class |

#### 3.1.1 Network Analysis

Although we use the contact matrices to model the connectivity of the campus, we can also use network analysis since we have access to individual-level data. In particular, we can build a network graph of every individual on campus and their connectivity to each other via classes only. From this network, we can determine characteristics of those individuals that have the highest rates of contact. These network analyses are presented in Appendix B.

To build the weighted undirected graph, we consider each individual affiliated with the university as a separate node. Edges are formed between two nodes if those two individuals share a class (either as students or as student/instructor). For each additional class individuals share, the edge weight is increased by one. Of course, the graph changes depending on whether all classes meet in person, or whether there is a class capacity. Supplementary Figure B1 (see Supplementary File 2) displays the resulting network for a full campus in which all classes meet in person. The network is colored by subpopulation (reddish purple corresponds to on-campus undergraduates, yellow represents off-campus undergraduates, sky blue for graduate students, and vermilion for faculty), the size of each node represents the weighted degree, and the edge thickness reflects weights. It is apparent that under normal conditions most of the campus is connected with one another. However, as seen in Supplementary Figure B4 (see Supplementary File 2), when classes with more than 25 students enrolled do not meet in person, much of the network becomes disconnected, i.e., having no contact with any other individuals.

With a resulting graph for the no intervention strategy that has over 9000 nodes and 1.6 million unique edges, it becomes necessary to use metrics to analyze exactly how the campus is connected. We report a histogram distribution of the weighted degree, a measurement calculating how many edges each node has. Histogram representations of the degree are displayed in Fig. 12. It is clear that capping in-person classes has a dramatic effect on the weighted degree, reducing maximal degree from 1,417 to 367. In fact, edges are reduced to *≈*0.13 million. Further, approximately 17% of campus individuals have 0 classroom contacts (no edges) when implementing a class cap strategy.

We can gather information about possible ‘super-spreaders’ by examining the individuals that have the highest degrees. Under the assumption of no interventions, it is clear the individuals with the highest degree are most often undergraduate students taking many introductory classes. For example, the individuals with the top 3 degree scores were all undergraduate students taking 4-5 lower level introductory classes (all living off-campus). When class caps are implemented, the structure of those with the highest interactions changes. Of the top 3 degree scores when imposing a class cap of 25, we have one lecturer teaching multiple labs and two graduate students that are taking a full load of classes and also teaching discussion sections and labs. This highlights how important it is to incorporate these subpopulations, who may serve as a vector for disease transmission, in our model.

### 3.2 Living Situations ℂ^*l*^

In addition to classroom contacts, most members of the community will also interact with other individuals based on their living situation. We assume that the living contacts are based on living situation only. Therefore, there is no mixing between subpopulations in our model. For example, on-campus undergraduate students do not live with off-campus undergraduate students and vice versa. This means that in our contact matrix (Table 2), living contacts exist only on the diagonals.

We assume that on-campus undergraduate students have contacts for approximately 10 hours/day with their direct roommate. To simulate the effects of encountering other individuals in their dorm, we assume that on-campus undergraduate students have 2 hours of contact per day multiplied by the average number of beds/bathroom. For example, if there are roughly 4 students per bathroom, they would experience 8 hours of additional contact per day.

The off-campus undergraduates are assumed to live with, on average, 3 off-campus undergraduate housemates. Since their living situation is likely larger than a dorm room, we assume less contact, at 4 hours/day contact with each roommate. Graduate students have a similar situation, except that we assume they live with 1.5 other graduate housemates. Similarly we assume 4 hours of contact per day with each roommate for each graduate student. We assume that faculty and staff to do not live with other faculty and staff.

### 3.3 Contact with Outside Community ℂ^*o*^

One of the most important aspects about a bubble-like community, from an infectious disease perspective, is that some key individuals have contact with the “outside community”. This is often overlooked in mathematical models, partly because the populations that interact with the outside world tend to be outnumbered by those contained fully in the bubble. However, these outside contacts cannot be ignored because they represent the potential for infection to infiltrate the “closed” community. These contacts are present in the fifth column of the contact matrix displayed in Table 2.

There are varying levels of contact with the surrounding community depending on which subpopulation a person is part of. We assume that there is little contact with the outside world if you live on-campus (1 hour of contact/day). For off-campus undergraduate students and graduate students, the number of contacts is higher due to increased shopping, transportation, etc., at 5 hours of contact/day. We assume that faculty and staff have the highest amount of contact with the outside community since many faculty live with families that are not affiliated with the university (15 hours of contact/day). However, as these numbers are not directly produced from known data, we also include a parameter *c*, a multiplier in front of the community contact matrix, that we vary.

### 3.4 Unscheduled Social Interactions ℂ^*s*^

One aspect of contact that has not yet been addressed is contact that occurs outside the classroom and living situation. In particular, we consider the effect of “unscheduled” social interaction in which members of the undergraduate student population meet for gatherings, unmasked, on a daily basis. Examples of these daily social interactions might include eating dinner with friends in the dining hall or forming an in-person study group for a course. In our contact matrix (Table 2), these are included in the undergraduate student populations (both on and off campus).

We incorporate unscheduled social interactions in our model in two ways: first the daily weekday interactions described above and secondly an increase in social interaction during the weekend. To calculate the daily social interactions, we scale the classroom contacts by 30%. Our assumption is that the amount of contacts each student has socially will be similar to those experienced in the classroom, especially as some of our “unscheduled” social interactions include course study groups. The scaling value is chosen as studies show that the contact-hours in ‘other/social’ is roughly 30% of the ‘school’ contacts [36, 37]. We assume that as class caps are implemented, unscheduled social contacts similarly decrease. Since many of these unscheduled social contacts are based on study groups arising from courses, unscheduled social contacts decrease as class caps are implemented. Thus, the social contact matrix ℂ^*s*^ is always 30% of the classroom contact matrix ℂ^*c*^. There is large uncertainty in number of social contacts, and so we multiply this contact matrix by the parameter *p*, which varies from 50% to 150% as shown in Table 1 as part of the sensitivity analysis. For the increased social interaction over the weekend, we multiply these daily social contacts by the parameter *w*, to simulate going to a larger gathering. This parameter and larger number of contacts is active from 5pm Friday until 5pm Sunday.

## 4 Results: Global Sensitivity Analysis

We are interested in the sensitivity of the cumulative number of infections at a point in time since the semester began, *C*(*t*_*i*_), and the sensitivity of the infection doubling time, Δ*T*, to the model parameters and initial conditions. We refer to initial conditions and model parameters as “model factors” in this text. We performed a global sensitivity analysis following [27] and estimated the first-order sensitivity index and the total-order effect through numerical model solutions generated by sampling from the input factor space, assuming that all parameters were uniformly distributed in their given range listed in Table 1.

### 4.1 Variance in Cumulative Infections and Infection Doubling Time

We first study the variance of the following model metrics: the time-varying cumulative infections, the doubling time, and number of the cumulative infections at the end of the fifteen-week term. As mentioned above, we do this by employing a global sensitivity analysis approach where we vary parameters independently and uniformly over their ranges. In our analysis we consider the quantifiable effect of three classes of model factors: infection parameters (*β, σ, φ, κ*^*R*^, *κ*^*V*^), contact parameters (*M, c, ω, p, m, α*), and initial conditions (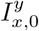 where *y* ∈ {*s, a*} and *x* ∈ {*d, u, g, f*}) (see Table 1 and Appendix A for model factor ranges and details).

Fig. 3 presents the behavior of the cumulative number of infections as a consequence of class caps (each column represents a different class cap: none, 100 students and 50 students) and vaccination status of the campus at the beginning of the term (each row). For simplicity, we assume that all undergraduates have the same vaccination fraction *v*_*u*,0_ = *v*_*d*,0_ and that graduate students and faculty have the same vaccination fraction *v*_*f*,0_ = *v*_*g*,0_, at the start of the semester. We consider the following three vaccination scenarios, Low: *v*_*u*,0_ = *v*_*d*,0_ = 0%, *v*_*f*,0_ = *v*_*g*,0_ = 0%; Medium: *v*_*u*,0_ = *v*_*d*,0_ = 40%, *v*_*f*,0_ = *v*_*g*,0_ = 50%; and High: *v*_*u*,0_ = *v*_*d*,0_ = 80%, *v*_*f*,0_ = *v*_*g*,0_ = 100%. The black line signifies the mean cumulative infections over time, while the pink and blue shadings show one and two standard deviations from the mean, respectively. The mean and coefficient of variation for the doubling time and total cumulative infections are reported in each subplot.

Without vaccination or class caps (Fig. 3(a)), we expect about 1,500 infections by the end of the semester (the baseline number of cumulative infections). By implementing a class cap of 50 students, the expected cumulative infections drops to about 200, an 86.8% reduction in cumulative infections (RECI) from the baseline (Fig. 3(c)). On the other hand, increasing vaccination rates to 100% for faculty and graduate students and 80% for undergraduate students reduces the expected cumulative infections to about 112 (92.6% RECI) by the end of the semester (Fig. 3(g)). With a class cap of 50 and high vaccination rates, we expect about 82 cumulative infections (94.6% RECI) by the end of the semester (Fig. 3(i)).

**Figure 3:**
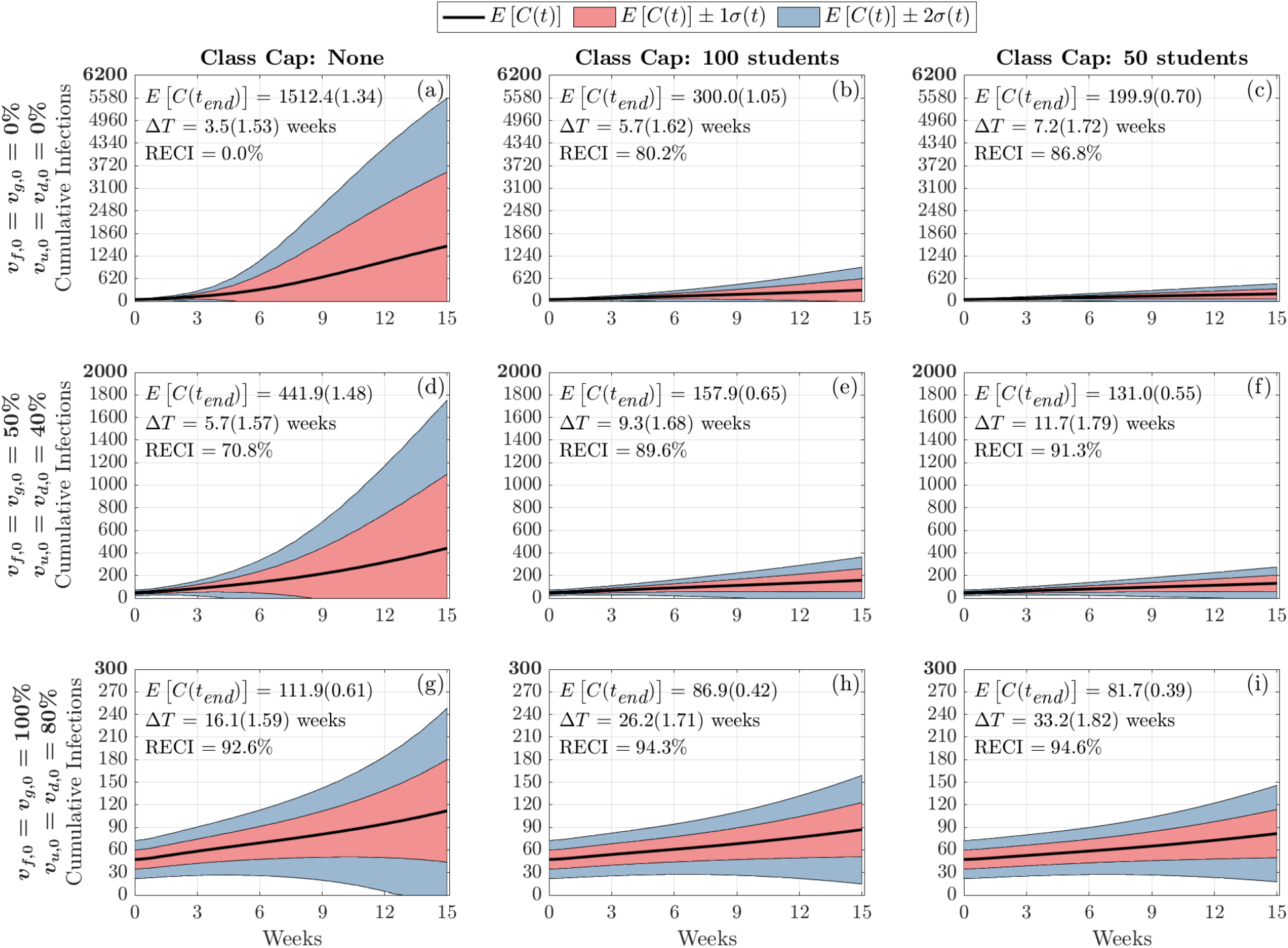
Distribution of Cumulative Infections. Figures show the distribution of cumulative infections over the span of a semester (15 weeks) and we allow the contact, infection parameters, and initial number of infectious individuals to vary (See Table 1). The mean and coefficient of variation for the doubling time and total cumulative infections are reported in each subplot. The reduction in expected cumulative infections (RECI) from case (a) with no vaccination and no class caps is presented in each subsequent case

In addition to examining the cumulative number of infections, we can also investigate how vaccination and class caps impact the doubling times of infections among the campus populations. Fig. 4 displays the infection doubling time in weeks (top row) and cumulative infections at the end of the semester (bottom row) as a function of vaccinated undergraduate students, *v*_*u*,0_ = *v*_*d*,0_. The columns correspond to faculty and graduate student vaccination rates 0% (left), 50% (middle), and 100% (right). Within each panel, the effects of incorporating class caps are portrayed with no class cap (yellow/thick line), 100-person cap (pink/medium line), and 50-person cap (teal/thin line). As we increase undergraduate student vaccination rates, we observe the lengthening of infection doubling time and we note that infection doubling times often exceed the length of the semester (Fig. 4(a)-(c)). We also see that increasing faculty and graduate student vaccination rates is not as effective in reducing the number of cumulative infections as vaccinating the larger undergraduate student population. This can be seen in Fig. 4(d)-(f) in the similarity between the three panels where the faculty vaccination is increased by 50% from left to right. The curves representing the cumulative infections, as a function of undergraduate vaccination rate, indicate a minimal change in the number of cumulative infections as faculty and graduate student vaccination rate is increased to 100%. This is likely because the faculty and graduate students only make up about 12% of the campus population.

It is apparent that the NPI of having large enrollment classes be remote drastically reduces the number of cumulative infections by the end of the semester, especially when the campus population is not significantly vaccinated (Fig. 4(d)-(f)). In the case when none of the population was vaccinated, by capping classes at 50 students, there is an 86.8% reduction in the expected cumulative infections by the end of the semester (Fig. 3(c)). Increasing the percentage of the vaccinated population also has a large effect in reducing the cumulative infections by the end of the semester. In the case when there is no class cap, having 80% of undergraduates and 100% of faculty, staff, and graduate students vaccinated resulted in a 92.6% reduction in the expected cumulative infections by the end of the semester (Fig. 3(g)). We observe from Fig. 4, it appears that under low vaccination, class caps have a higher effect at reducing the number of cumulative infections. However, under high vaccination, implementing class caps has less of an effect in reducing the expected number of cumulative infections by the end of the semester.

**Figure 4:**
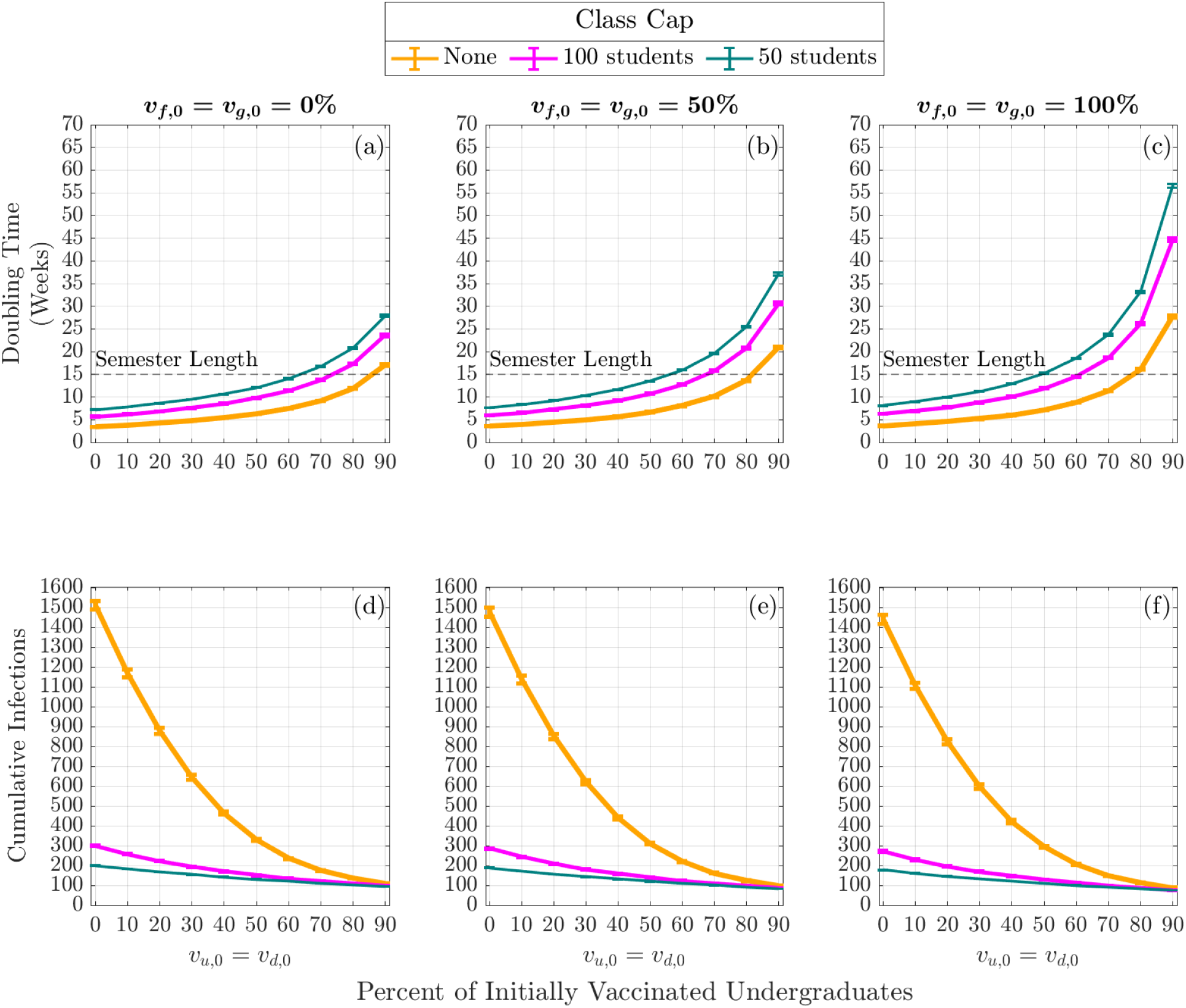
Expected Infection Doubling Times and Cumulative Infections by Class Capacity and Percent of Vaccinated Undergraduates. The expected cumulative number of infection (cumulative infections) by the end of the semester and the expected doubling time computed during the first four weeks of the semester. The error bars are a 95% confidence interval

### 4.2 Sobol Analysis of the Variance in Cumulative Infections and Infection Doubling Time

The first-order Sobol sensitivity index 𝕊_*i*_ measures the direct effect that each model factor *θ*_*i*_ ∈ (*β, σ, φ, κ*^*R*^, *κ*^*V*^, *M, c, ω, p, m, α*, 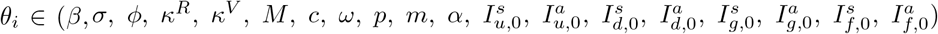) has on the variance of the model output. The total-order Sobol sensitivity index 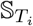 measures the total effect (direct and through interactions with other model factors) each model factor, *θ*_*i*_, has on the variance of the model output. As mentioned above, we consider the sensitivity of two model outputs: the cumulative number of infections over time and the doubling time of the infection. We categorize the parameters for the sensitivity analysis into three groups: infection parameters (*β, σ, φ, κ*^*R*^, *κ*^*V*^), contact parameters (*M, c, ω, p, m, α*), and initial conditions 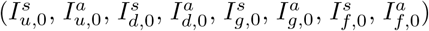. In each sensitivity analysis figure, we show 9 subfigures illustrating three class cap scenarios—no class cap (the first column), class cap with 100 students (the second column), and class cap with 50 students (the third column)—along with three vaccination scenarios—(i) 0% vaccination (the top row); (ii) 50% of faculty, 50% graduate students, and 40% of undergraduate students vaccinated (the middle row); and (iii) 100% of faculty, 100% graduate students, and 80% of undergraduate students vaccinated (the bottom row).

We note that the initial conditions do not contribute to the sensitivity of either of our metrics in a fashion that is dependent on the vaccination or NPIs. As such, we include those figures in Appendix C.

#### 4.2.1 Sensitivity of Infection Doubling Time

We now turn to examine the global sensitivity analysis with respect to infection doubling time (Δ*T*). Fig. 5 displays the global sensitivity analysis of infection doubling time for infection and contact model parameters, while Fig. 13 presents the global sensitivity analysis of infection doubling time with respect to initial conditions.

When examining Fig. 5, we can see that across these vaccination scenarios and class caps, infection transmission rate *β* is the most significant parameter having the greatest individual effect on the variance in infection doubling time (Δ*T*). Across these scenarios, transmission rate, *β*, captures 40% to 60% of the variance in first-order effects, while considering interaction effects with other model factors (the total-order effect), *β* captures 40% to 70% of the variance in infection doubling time. That is, *β* has the largest effect on infection doubling time Δ*T* among all model factors. We also see that both greater vaccination rates and lower class caps lead to a decrease in the effect of transmission rate on infection doubling time. The strong sensitivity of infection doubling time on transmission rate implies that anything that can be done to lower the transmission rate, such as wearing masks or improving HVAC systems, can have a large impact on infection dynamics at the beginning of the semester.

Other parameters to which infection doubling time is sensitive to include contact parameters: infections from the outside community, *M*, community contact multiplier, *c*, weekend contact multiplier, *w*, social contact percentage, *p*, mask usage, *m*, and the probability that a symptomatic individual will self-isolate, *α*. From Fig. 5(a), under no class cap and no vaccination we observe an infection doubling time of Δ*T* = 3.5 weeks (the number of cumulative infections would double in 3.5 weeks) and the sensitivity of infection doubling time is similarly affected by transmission from the community (through *M* and *c*) and transmission on campus (through *p, w*, and *m*). The sensitivity from parameters of each type of contact, off-campus or on-campus contacts, capture about 10% of the variance in infection doubling time, however as the class cap is reduced to 50 students, Fig. 5(c), infection doubling time increases to Δ*T* = 7.2 weeks and the effect of mask, social, and weekend contact parameters is reduced. In this case, the effect of infections from the outside community, *M*, and community contact multiplier, *c*, become more influential. This indicates that when contacts are reduced on campus through NPIs that reduce the number of people that meet in one location, this reduces the effect from the transmission rate *β* and the social and weekend contacts, and infections from community and contact with the community have a greater effect on infection doubling time of cumulative on-campus infections. When vaccination is increased, as in Fig. 5(g), under no class caps, with faculty and graduate student vaccination rates of 100%, and 80% of the undergraduate student population vaccinated, at the beginning of the term, we see an increase in infection doubling time to Δ*T* = 16.1 weeks (longer than our 15 week term) but no real qualitative change in the sensitivity of model factors when compared with reducing class cap. This minimal change in sensitivity to parameter values is because vaccination reduces the number of infected and susceptible individuals, but vaccination does not reduce contacts in the way that implementing class caps does by directly reducing person to person interactions. This analysis highlights the importance of including community interactions in models of bubble-like communities. Moreover, this shows that an effort to reduce transmission on campus will require attention to infection rates in the surrounding community.

Across scenarios in Fig. 5, the probability that a symptomatic individual will self-isolate, *α*, and the amount of time spent in the ‘exposed class’, *σ*, have non-zero total-order effect and no first-order effect. This indicates that *σ* and *α* alone do not contribute significantly to the model variance, but through interaction with other parameters can have a significant impact on infection doubling time. Unfortunately, determining exactly which model factors *σ* and *α* are interacting with, and to what extent, is not computationally tractable with this Sobol analysis. Another parameter that has a non-zero effect on doubling time is *m* or mask usage, where 1 − *m* is the reduction in infection transmission probability by wearing a mask. From Fig. 5 we observe that as we increase vaccination rates (moving down a column), we can see that the impact of wearing a mask, model factor *m*, stays rather consistent in its importance. This indicates that masks are still crucial for controlling the spread of disease even as vaccination rates increase. However, when decreasing the class caps, we can see that the sensitivity to *m* decreases. Thus, NPIs such as wearing masks and reducing contact by moving courses online are still important in controlling the number of infections on campus. The duration of naturally acquired immunity 1*/κ*^*R*^ and immunity through vaccination 1*/κ*^*V*^, do not affect infection doubling time since both types of immunity are assumed to last longer than three months (See Table 1) and infection doubling time is computed using cumulative infections observed within the first month.

**Figure 5:**
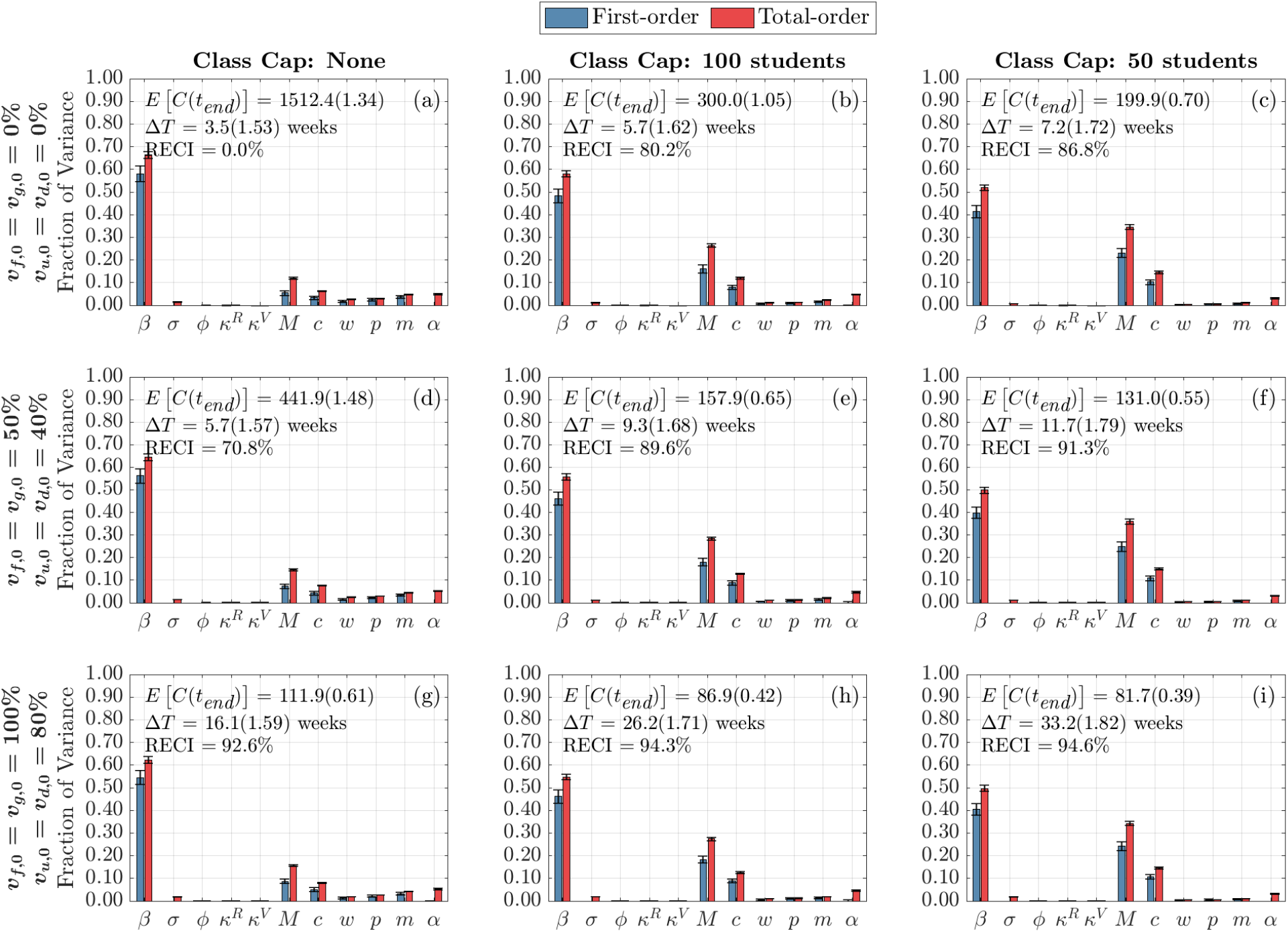
Global Sensitivity Analysis of Infection and Contact Parameters on infection Doubling Time. First-order (blue) and total-order (red) Sobol indices are shown as well as the standard errors. The mean and coefficient of variation for the doubling time and total cumulative infections are reported in each subplot. Each column represents three class cap scenarios: none, 100 student, and 50 student caps. Each row represents one of three vaccination scenarios at the start of the semester. First row: 0% vaccination; second row: 50% of faculty, 50% graduate students, and 40% of undergraduate students vaccinated; third row: 100% of faculty, 100% graduate students, and 80% of undergraduate students vaccinated

Fig. 13 displays the sensitivity of infection doubling time to the number of initially infectious individuals on campus. The distribution and number of initially-infectious individuals at the start of the semester (undergraduate students, graduate students, and faculty and staff) do not play a large role in determining infection doubling time. This is intuitive, since infection doubling time measures the time it takes to *double* the number of cumulative infections and should be the same whether we start with one infectious individual or 100 infectious individuals (See Fig. 2).

#### 4.2.2 Sensitivity of Cumulative Infections at End of Term

Here, we are interested in quantifying the effect of each parameter on the cumulative number of infections at the end of the term for each vaccination and class cap scenario. Fig. 6 displays the global sensitivity analysis for the cumulative infections at the end of the semester with respect to infection and contact parameters. We find that as vaccination rates increase and class caps are implemented, the most sensitive parameters begin to change. Under the scenario with no vaccinations and no class caps (Fig. 6(a)), the most important parameters are the transmission rate, *β*, weekend contact multiplier, *w*, social contact percentage, *p*, mask usage, *m*, and the probability that a symptomatic individual decides to self-isolate, *α*. By incorporating different class caps, the landscape of sensitivity continues to change. The first-order effect of self-isolation, *α*, mask usage, *m*, social contact percentage, *p*, and weekend contact multiplier, *w* become less important, while the parameters governing the outside community start to play a larger role. The probability that Merced community individuals are infectious, *M*, and the community contact multiplier, *c*, start to influence the cumulative number of infections. This underscores the importance of educating local communities to help reduce the spread of infections in the community. As vaccination rates are increased, without class caps, we see a similar shift towards a decrease in first-order effects from mask usage, *m*, social contact percentage, *p*, and weekend contact multiplier *w*, however, these parameters remain influential through interaction effects with other parameters. When both vaccination and class caps are utilized, the community parameters become more important than the transmission rate, see Fig. 6(i).

**Figure 6:**
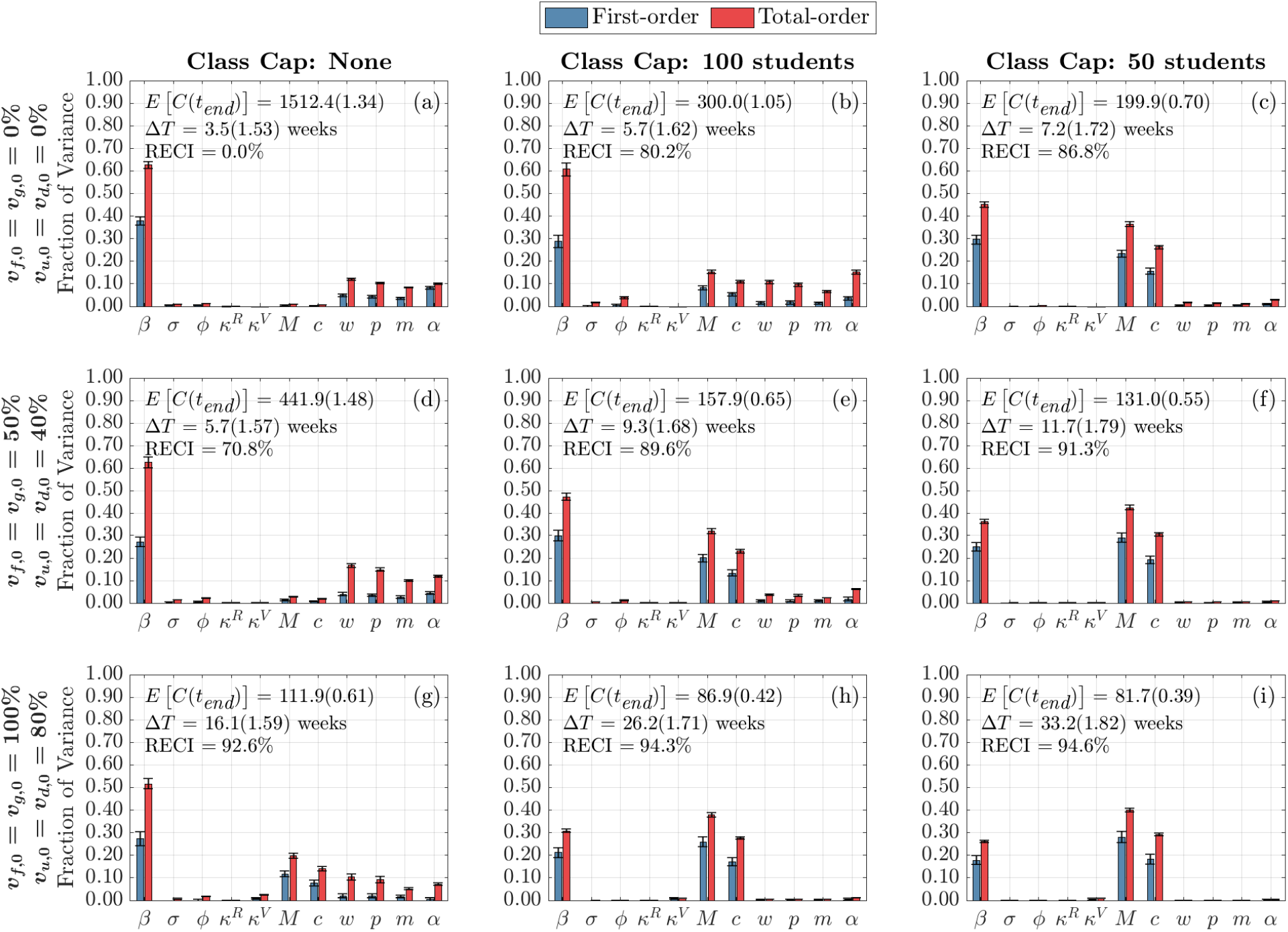
Global Sensitivity Analysis of Infection and Contact Parameters on Cumulative Infections at the End of the Term. First-order (blue) and total-order (red) Sobol indices are shown as well as the standard errors. The mean and coefficient of variation for the doubling time and total cumulative infections are reported in each subplot. Each column represents three class cap scenarios: none, 100 student, and 50 student caps. Each row represents one of three vaccination scenarios at the start of the semester. First row: 0% vaccination; second row: 50% of faculty, 50% graduate students, and 40% of undergraduate students vaccinated; third row: 100% of faculty, 100% graduate students, and 80% of undergraduate students vaccinated

We can also examine the sensitivity of cumulative infections to the initial number of infectious individuals on campus. Fig. 14 presents this sensitivity analysis. It is apparent that under no interventions, the initial number of infectious individuals has little effect on the cumulative number of infected individuals at the end of the term. Similarly, when incorporating class caps, the initial conditions are insensitive. However, under high vaccination rates (80% undergraduate students and 100% faculty, staff, and graduate students), the cumulative number of infections is slightly sensitive to the initially infectious individuals on campus – with slightly higher sensitivity to undergraduate student infections, both symptomatic and asymptomatic. The sensitivity in those scenarios is present because the final number of cumulative infections observed at the end of the term is mainly from those infected at the start of the semester and undergraduate students make up the greatest proportion of the campus population (about 88% of the on-campus population consists of undergraduates). This tells us that under the best-case scenario with both high vaccination and class caps, it is important to minimize the number of infectious students at the start of the term.

#### 4.2.3 Sensitivity of Time-Varying Cumulative Infections

In order to dissect the forces that contribute most strongly to the cumulative infections, we next consider their time-varying first and total-order Sobol indices. That is, we now look at the total contributions to the variance in cumulative infections over time. We separate the impact of infection parameters (Figs. 7 and 9), contact parameters (Figs. 8 and 10) and initial conditions (Figs. 15 and 16). Because the time-varying first and total order indices for initial conditions show that their effect is limited to the start of the term, and that their effect is greatest when the initially infected made up the majority of cumulative infections observed in one semester, we include those figures in Appendix C.

**Figure 7:**
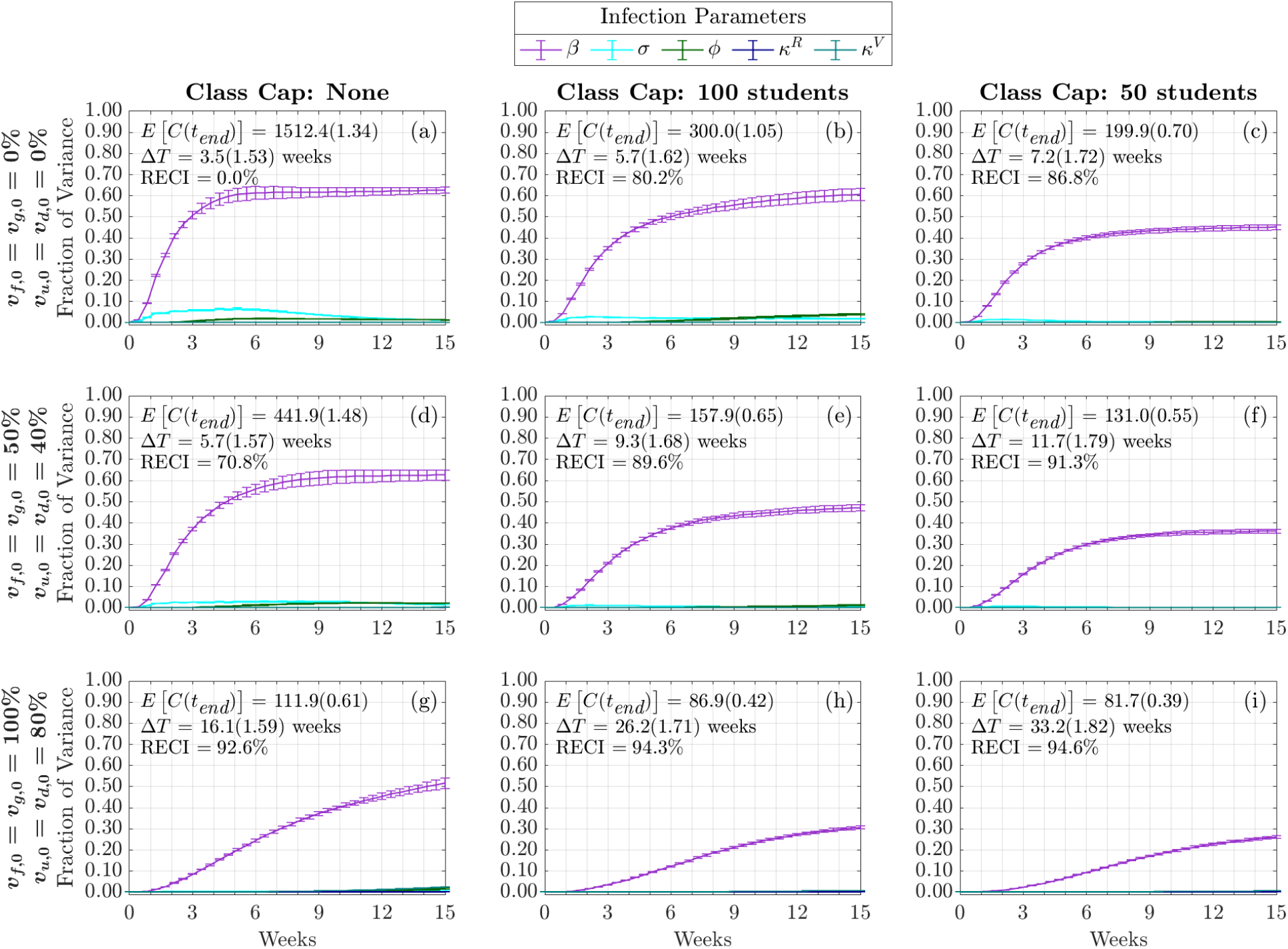
Time-Varying Total-Order Effect of Infection Parameters on Cumulative Infections. The total-order Sobol indices are shown as well as the standard errors. The mean and coefficient of variation for the doubling time and total cumulative infections are reported in each subplot. Each column represents three class cap scenarios: none, 100 student, and 50 student caps. Each row represents one of three vaccination scenarios at the start of the semester. First row: 0% vaccination; second row: 50% of faculty, 50% graduate students, and 40% of undergraduate students vaccinated; third row: 100% of faculty, 100% graduate students, and 80% of undergraduate students vaccinated

First, we consider the time varying impact of infection parameters on the cumulative infections. As shown in Figs. 7 and 9, regardless of the vaccination status or class cap scenario, the strongest contributor at all times comes from *β*. This is consistent with the results from Fig. 5, where we look at the sensitivity to the infection doubling time. We now see that this effect increases during the early part of the academic term. In the case of no vaccination and no class caps, Fig. 7(a), the amount of time spent in the ‘exposed state’, 1*/σ*, becomes influential early in the semester and reaches its strongest contribution to cumulative infections during the fourth week of the semester, the effect from *σ* then decreases. Both increasing vaccination and implementing class caps have the effect of delaying and dampening the contribution from *σ*. The probability of becoming asymptomatic, *φ*, at most explains about 5% of the variance in the case with no vaccination and a class cap of 100 students. Also, the duration of immunity from both vaccination, 1*/κ*^*V*^, and natural immunity, 1*/κ*^*R*^, do not affect the variance in cumulative infections, including through interactions with other parameters. This is expected since we are only modeling a 15-week term, and both natural and immunity through vaccination lasts at least 12 weeks (see Table 1).

Next, we present the role of contact parameters in Fig. 8 and Fig. 10. In Fig. 8, we note that importance of contact parameters depends not only on vaccination and class cap scenario, but also on time. In the case of no vaccination and no class cap, Fig. 8(a), we see that both the weekend contact multiplier, *w*, and social contact percentage, *p*, are initially the most influential among all the contact parameters but their effect decreases after the sixth week. We observe that as vaccination is increased without decreasing the class cap, Fig. 8(d) and Fig. 8(g), the effect on cumulative infections from these parameters is delayed towards later in the semester. However, as the person-to-person contacts on campus are reduced through class caps as in Fig. 8(b) and Fig. 8(c), we see the sensitivity to *w* and *p* replaced by the sensitivity to *M*, the percentage of infectious people in the community and *c*, the community contact multiplier. This is similar to the results from the sensitivity analysis in Figs. 5 and 6. In the case with no vaccination and no class cap, Fig. 8(a), we can see how *α*, the probability that a symptomatic person will self-isolate, becomes more influential over time but reaches a fixed effect of about 10% of the variance. Without class caps, as vaccination is increased to medium and high vaccination, Fig. 8(d) and Fig. 8(g) respectively, we see that the effect due to *α* is delayed to later in the semester but still reaches 10% of the variance by the end of the semester. However, under low vaccination and a class cap of 100 students (Fig. 8(b)), *α* becomes more significant, explaining more than 15% of the variance by the end of the semester. When comparing Fig. 8, the total-order effect, and Fig. 10, the first-order index, in terms of *α* we can see that most of the contribution to the model variance seen in Fig. 8 comes from the interaction effects with other model parameters. This means that in bubble-like communities with low vaccination rates and no class caps policy, making sure that students self-isolate if they are symptomatic plays a large role in affecting the cumulative number of infections observed over the course of the semester.

**Figure 8:**
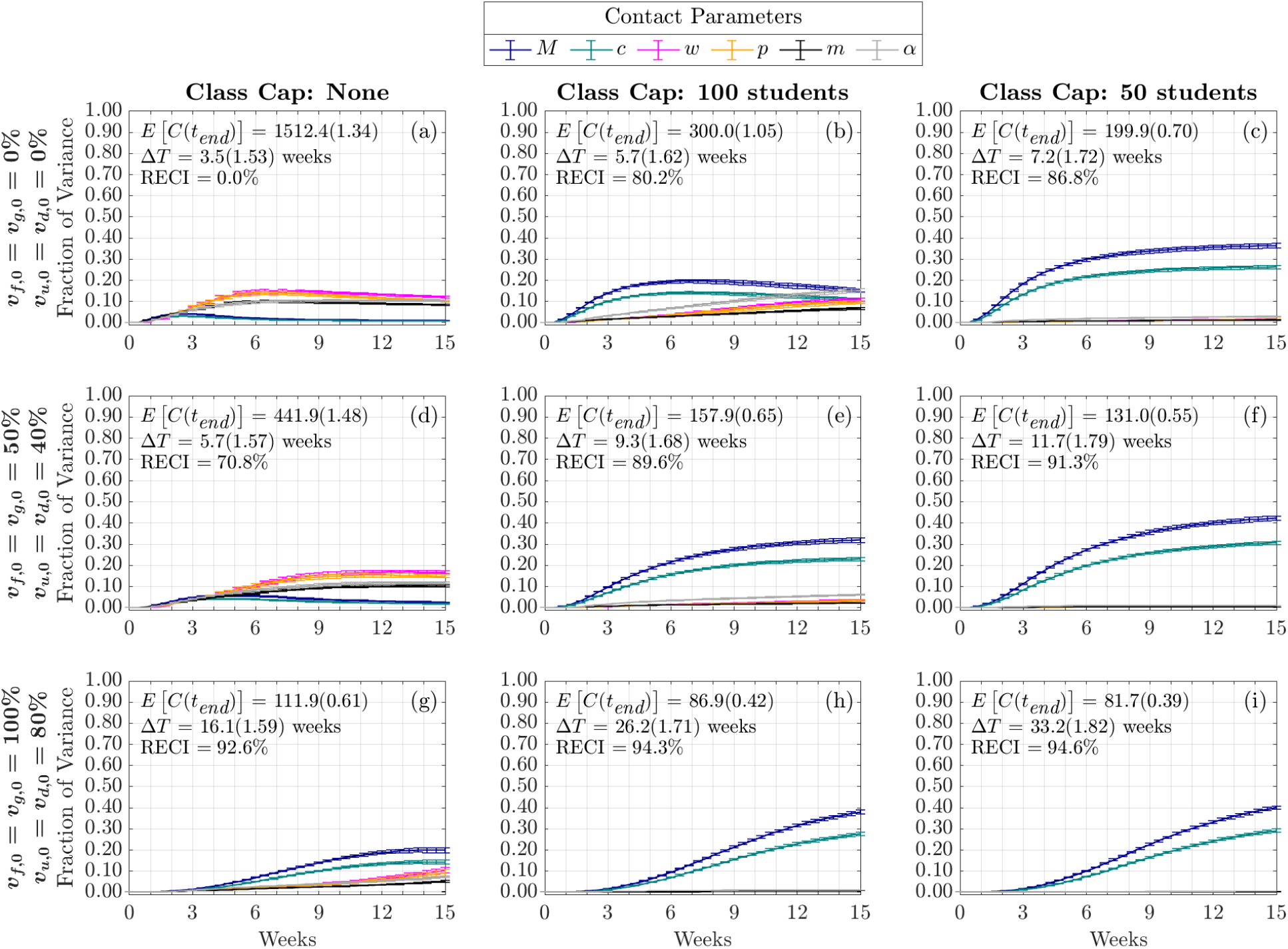
Time-Varying Total-Order Effect of Contact Parameters on Cumulative Infections. Each column represents three class cap scenarios: none, 100 student, and 50 student caps. Each row represents one of three vaccination scenarios at the start of the semester. First row: 0% vaccination; second row: 50% of faculty, 50% graduate students, and 40% of undergraduate students vaccinated; third row: 100% of faculty, 100% graduate students, and 80% of undergraduate students vaccinated

From Figs. 15 and 16 we see that the infectious people at the beginning of the term only affect the cumulative infection within the first few weeks of the term. While we do observe different levels of sensitivity to these initial conditions, these effects are separated by population size. Among those that are infectious at the start of the term, those that are symptomatic have a slightly larger effect than those that are asymptomatic even though asymptomatic people are 50% less infectious than symptomatic people in our model. The largest effect on cumulative infections, from up to 1% of the population that is initially infectious, comes from undergraduate students that live off-campus (5,441 students), followed by students living in the dorms (2,885 students) and the least influential infections, consistently explaining less than 1% of the variance, are both graduate student (721 graduate students) and faculty/staff initially infected (424 faculty & staff). In all cases, we see that both increased vaccination and implementing lower class caps extend how far into the term these model factors remain significant. For example, in Fig. 15(i) and Fig. 16(i), we see that the effect from the initially infectious undergraduate populations persists until the end of the term. That is because high vaccination and smaller class caps leads to fewer cumulative infections by the end of the term, and the initial infection make up a large number of the cumulative infections observed over the term.

For non-linear models, it is not generally true that the first and total order Sobol indices will be equal. This only happens in cases where the model factors only have a linear contribution to the model variance as in the case of the initial conditions (Figs. 15 and 16). However, in this case we note that the importance of model factors (i.e., the rank of their contribution to the variance) is similar for first order and total order time varying Sobol indices (compare Fig. 7 to Fig. 9 and Fig. 8 to Fig. 10). However, there are a few intriguing differences to point out. Most notably, the first order index for *β* (Fig. 9(a)) may exhibit an internal peak rather than simply increase to saturation. Our interpretation again would involve a change in the dynamics from early in the semester with a large susceptible population to later in the semester with less susceptible people that can become infected. For the case with no class cap and no vaccination, the transmission rate *β* is the parameter that has the most significant direct effect on the output variance (Fig. 9(a) and Fig. 10(a)). This indicates that any interventions that can lower the transmission rate, *β*, would significantly impede the transmission. As shown in Fig. 9(a), we observe that the fraction of variance due to *β* begins to decrease after week 3 and begins increasing again after week 6. From Fig. 7(a) of the time varying total-order effect, the saturation in *β* is likely due to the increasing interaction effects with the contact parameters (Fig. 8), and with the other model factors that have a non-zero effect.

**Figure 9:**
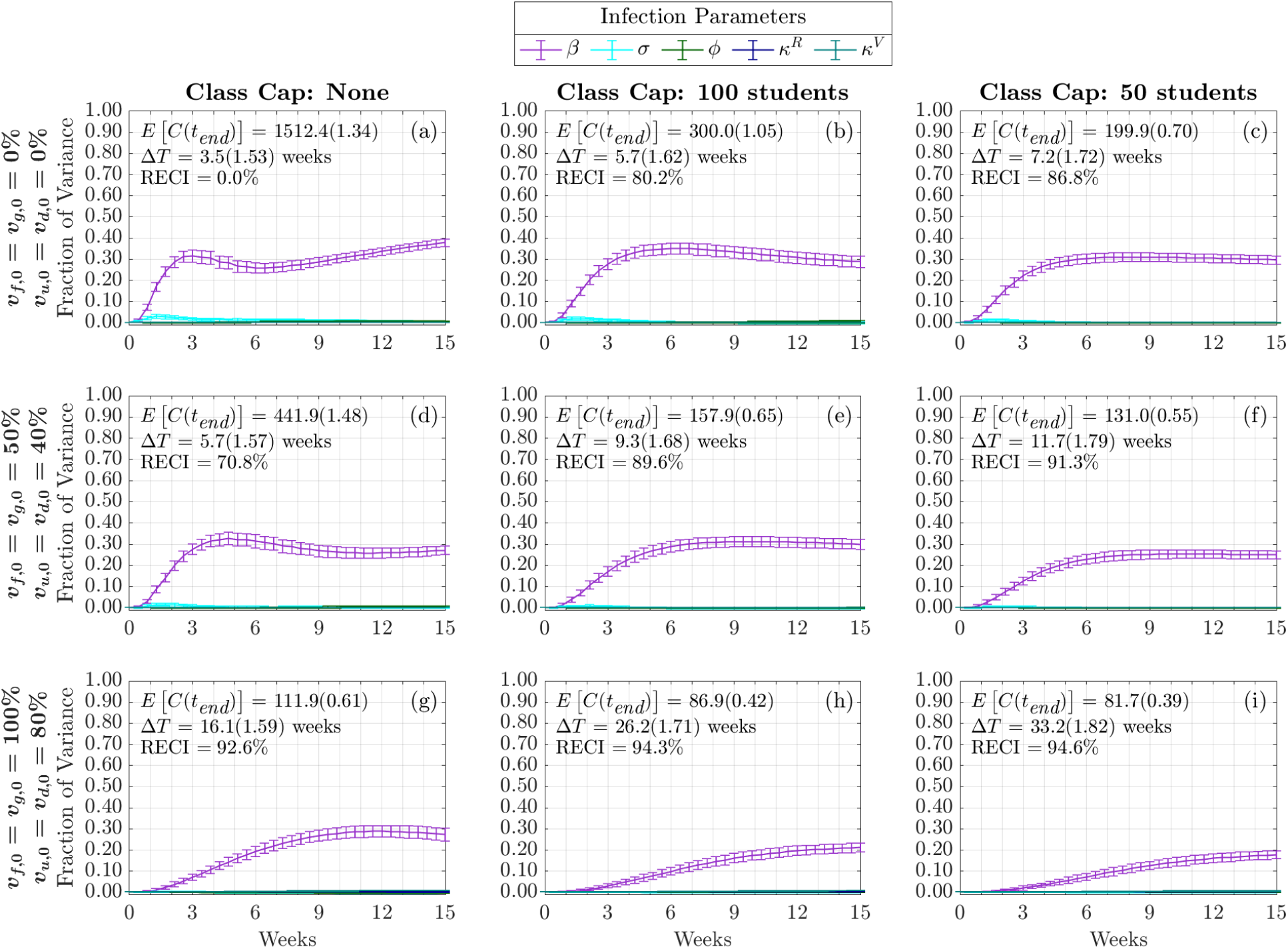
Time-Varying First-Order Effect of Infection Parameters on Cumulative Infections. Each column represents three class cap scenarios: none, 100 student, and 50 student caps. Each row represents one of three vaccination scenarios at the start of the semester. First row: 0% vaccination; second row: 50% of faculty, 50% graduate students, and 40% of undergraduate students vaccinated; third row: 100% of faculty, 100% graduate students, and 80% of undergraduate students vaccinated

## 5 Discussion and Conclusion

In this manuscript, we introduced an ODE-based SEIR model with multiple subpopulations (undergraduate students living on-campus, undergraduate students living off-campus, graduate students, and faculty and staff) on a campus that interacts with the outside community. We discussed how to use registrar information to estimate contact-hours due to classes. We examined the effects of NPIs such as social distancing in the form of transitioning large classes to an online format, mask usage, and considered differing vaccination rates with waning immunity for both natural immunity and immunity from vaccination among the different campus subpopulations. We acknowledge that there are other methods of incorporating heterogeneous interactions, such as agent-based modeling and stochastic modeling [38, 39]. Our proposed model preserves some aspect of heterogeneity while remaining open to traditional methods of analysis and being computationally inexpensive. This allowed us to evaluate different intervention strategies and obtain rapid results to assist the administration in making decisions regarding the mitigation of COVID-19 on the university campus.

**Figure 10:**
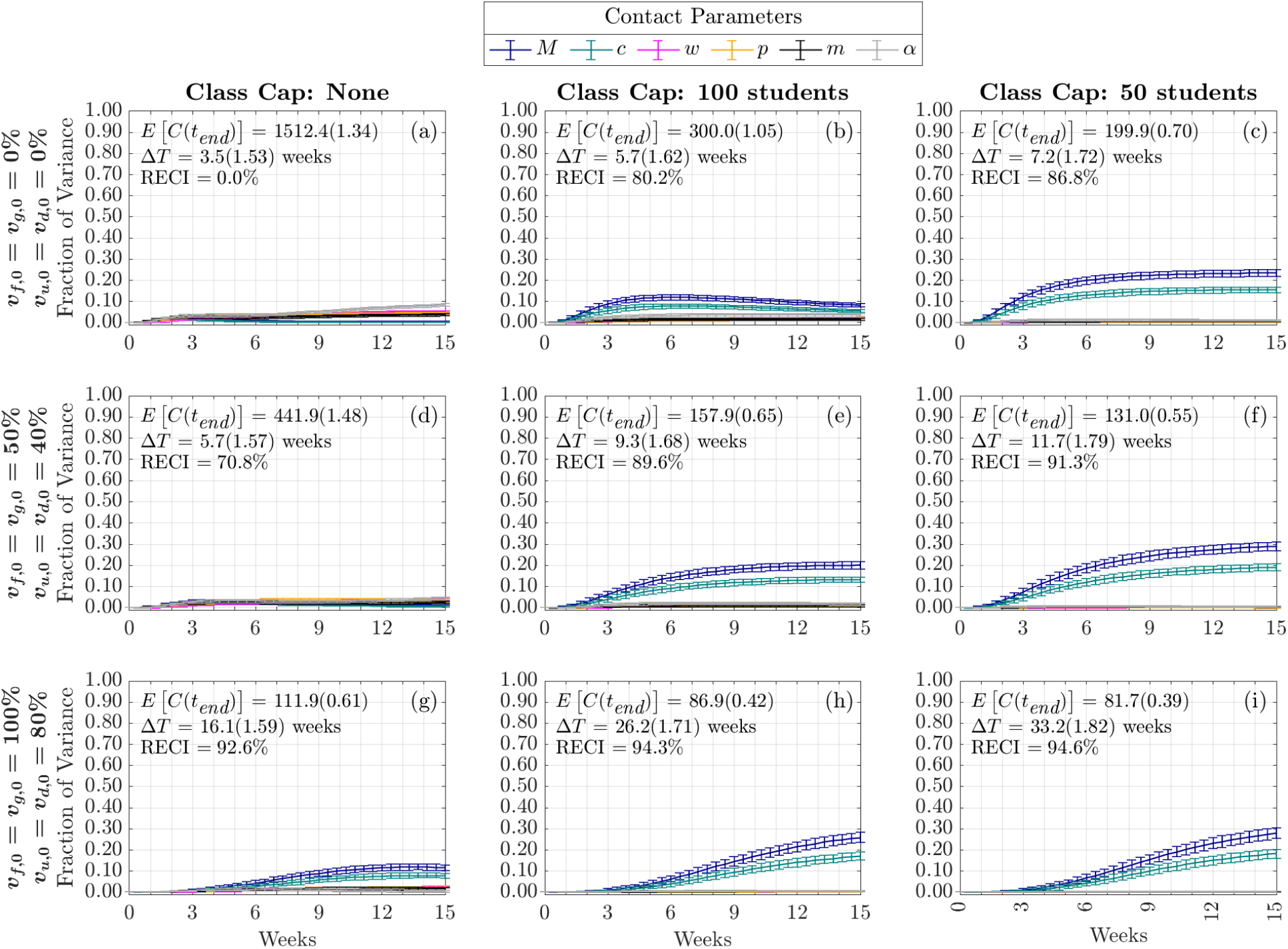
Time-Varying First-Order Effect of Contact Parameters on Cumulative Infections. Each column represents three class cap scenarios: none, 100 student, and 50 student caps. Each row represents one of three vaccination scenarios at the start of the semester. First row: 0% vaccination; second row: 50% of faculty, 50% graduate students, and 40% of undergraduate students vaccinated; third row: 100% of faculty, 100% graduate students, and 80% of undergraduate students vaccinated

We examined the sensitivity of two epidemic characteristics, the infection doubling time and the cumulative number of infections over the course of the semester, to 19 model factors that fall into three categories: infection parameters, contact parameters, and initial conditions, under varying levels of vaccination rates and class caps. Through our analysis, we found that implementing class caps (or limiting the number of people that can meet in an enclosed space) and greater vaccination rates, led to changes in the rank importance of the model factors in their contribution to the model variance. We found that, in scenarios with increased vaccination rates without class caps, the social contact multiplier, weekend contact multiplier, and probability to self-isolation when symptomatic are the most significant model factors contributing to the infection doubling time and the number of cumulative infections observed over the course of one 15-week academic term (one semester). However, when decreasing person-to-person contacts through smaller class caps, under low vaccination, the contacts with infectious people from the outside community play a predominant role in affecting the number of cumulative infections for the campus population and directly affects the infection doubling time. This work highlights that even when the campus implements and regulates NPIs, or can require vaccination, the outside community still have a large effect on the infections observed on campus. Therefore, it will be necessary for universities to work with their surrounding communities to help limit the spread of COVID-19.

Many of the parameters, such as the contact matrices, were directly informed from campus data. Thus, it may be that these results may not hold for other campuses that differ from our own (e.g., no large classes, campuses that are more integrated with the surrounding community, or campuses in urban settings). However, our flexible framework allows other universities to alter parameters and calculate contact matrices from their registrar data in order to make decisions about intervention strategies and to perform sensitivity analyses for their own campuses.

The parameters that were not directly calculated from data remain a source of uncertainty in the model. Although many of the parameter values were taken from literature values, the accuracy of those estimates is unknown. Therefore, model validation with real data remains a future direction for exploration. We also plan to replace the way that we model infections originating from the surrounding community, which is currently a constant, with dynamic data from county dashboard reports. We can also study the impact of a surge in COVID-19 infections in the off-campus community on the campus population by introducing a pulse in the community parameter.

We note there are several limitations with the current study. In our model it is assumed that vaccination makes the recipient 100% immune with the possibility of breakthrough cases through waning immunity. A logical future direction would include booster shots and the introduction of novel variants (e.g., delta, omicron). This will be especially important when modeling an academic year as opposed to one 15-week academic term. Another modeling option would be to include vaccinated subpopulations with decreased transmission rates and the possibility of shorter convalescence times. As differing variants have become dominant, the parameters would need to be updated to reflect appropriate transmission rates and, perhaps, a shortened infectious period of time. We also assumed 100% compliance with interventions, meaning that every member of the campus community wears well-fitting masks at all times when they are indoors. In our work, we attempted to capture the varying degrees of mask usage and effectiveness by varying the mask efficiency from having no effect to reducing transmission by up to 60%.

Overall, our model exhibits results consistent with current public health messaging. Our model suggests that, despite increasing vaccination rates, it remains important to continue to socially distance, to wear masks, and to implement class caps to reduce the transmission of COVID-19. It is likely that these results will not hold as more COVID-19 variants emerge, however our modeling framework can be readily adapted to account for such novel infections.

## Data Availability

Open-source code for the global sensitivity analysis and contact matrices can be found at https://github.com/FS-CodeBase/gsa_of_covid19_transmission_on_a_university_campus.

## Acknowledgments

The work presented in this manuscript is based upon work supported by the National Science Foundation (DMS–1840265 and ACI-1429783), the Joint Initiative to Support Research at the Interface of the Biological and Mathematical Sciences between the National Science Foundation Division of Mathematical Sciences (NSF-DMS) and National Institutes of Health National Institute of General Medical Sciences (NIH-NIGMS) (Grant No. R01-GM126548), and UC Merced COVID-19 Seed Grant.

We would like to acknowledge Drs. Folashade Agusto, Joan Ponce, and Omayra Ortega for detailed discussions in formulating the original version of the UC Merced model. We also acknowledge the guidance and support from Dr. Thelma Hurd, Director of Medical Education, Professor of Public Health, who was the head of UC Merced’s COVID Task Force.

## A Details of the Model

We track infection and immune status by modeling four campus subpopulations each with eight compartments. A schematic is provided in Fig. 1(b) for a single subpopulation, undergraduate students who live off-campus (*u*). Fig. 11 here displays all the campus subpopulations, undergraduate students who live off-campus (*u*), undergraduate students who live on-campus in the dorms (*d*), graduate students (*g*), and faculty and staff (*f*) as they progress through different stages of the COVID-19 infection.

Next, we discuss in more details the parameterization of our ODE model, the parameters are arranged in the same order as in Table 1. *β*: transmission rate of symptomatic infectious individuals per contact-hour, (5.74–24.86)×10^−4^.

The review by [40] compared 12 studies with the mean of *R*_0_ estimates ranging from 1.5 to 6.68 with a median of 2.79. On the other hand, 13 out of 20 estimated values of *R*_0_ listed in the review [41] were between 1 and 3. Although [42] estimated a median *R*_0_ to be 5.9 in the United States during early COVID-19 pandemic, their inference was driven by data collected from highly populated areas such as New York city. Merced, CA is a mid-sized rural city, and the UC Merced campus population is younger and healthier and has better access to vaccines and healthcare than the broader population. Thus we choose the ranges of *β* corresponding to *R*_0_ ∈ [1.5, 6.5] (see Table 1) to capture the transmission rates associated with a relatively wide range of population densities.

1/*σ*: duration in exposed state, 1–7.5 days.

The incubation period for the original strain of COVID-19 is estimated to be 4 to 6 days [43, 44]. Previous mathematical modeling efforts have used incubation period as a proxy for duration in exposed state [45]. However, we note that since incubation time represents the time between exposure and symptom onset and we are modeling the time between exposure and *infectiousness*, we subtract the one to two day pre-symptomatic infectious period [46]. This gives a baseline value around 3-5 days, which is then varied from 1 to 7.5 days for the sensitivity analysis.

*φ*: probability that an exposed individual will become asymptomatic, 20%–80%.

*ζ*: fraction of *β* for asymptomatic individuals, 50%.

Currently there is limited data on the full characterization of asymptomatic SARS-CoV-2 infection—such as the proportion of asymptomatic infection among the general population and the subpopulation with confirmed infection, and the transmission rate of asymptomatic individuals compared to symptomatic individuals—given that COVID-19 surveillance systems rely on symptom-based screening primarily. On the other hand, many studies do not distinguish presymptomatic infection (no symptoms at screening point but develop symptoms later on) and truly asymptomatic infection (no symptoms at screening point and never develop any symptoms) very well, i.e., the imprecise use of the term “asymptomatic”.

A systematic review [47] analyzed 14 longitudinal studies and estimated a median of 72.3% (IQR 56.7%-89.7%) of individuals who tested positive but had no symptoms at the time of testing would remain asymptomatic. Another systematic review [48] estimated a median of 35.1% (95% CI: 30.7%-39.9%) of laboratory-confirmed cases were truly asymptomatic by analyzing 170 studies, they observed greater asymptomaticity in children than the elderly, and lower asymptomaticity among cases with comorbidities than cases with no underlying medical conditions. A meta-analysis of 130 studies [49] found an interquartile range of 14% to 50% of people infected with COVID-19 that was persistently asymptomatic. To account for the large uncertainty, we choose to vary *φ* from 20% to 80% in our global sensitivity analysis.

Using mobility data and case reports within China and Bayesian inference along with a mathematical model, [50] estimated that a median of 86% of infections went undocumented and these undocumented infections were 55% (95% CI 46-62%) as contagious as documented infections. A systematic review [49] suggests that asymptomatic infection might be less contagious than symptomatic infection (risk ratio 0.32 with 95% CI 0.16-0.64). For simplicity, we choose *ζ* to be 0.5.

1/*κ*^*R*^: average duration of naturally acquired immunity, 90–270 days.

1/*κ*^*V*^ : average duration of vaccine-mediated immunity, 180–270 days.

Long-term longitudinal studies on COVID-19 reinfection can provide crucial data that would enhance our understanding of the durability of naturally acquired and vaccine induced immunity. However, designing such studies and interpretation of results is very complex due to the emergence of variants, the potential of hybrid immunity from both infection and vaccine, different vaccine options and dosing schedules, and heterogeneity in populations. We choose those lower bounds for 1/*κ*^*R*^ (90) and 1/*κ*^*V*^ (180) based on the 5% quantile of the predicted time to breakthrough infection following mRNA vaccination (3.5 months after vaccination with BNT162b2 or mRNA-1273) and viral vector vaccines (4.3 and 2.6 months after vaccination with ChAdOxa and Ad26.COV2.S, respectively), and the 5% quantile of the predicted time to reinfection following natural infection (3.5 months) in [51]. We set the upper bounds for both 1/*κ*^*R*^ and 1/*κ*^*V*^ to be 270, longer than the 15-week semester period.

*M*: probability that outside (Merced) community individuals are (asymptomatically and symptomatically) infectious, 0%–1%.

*ψ*: probability that outside (Merced) community individuals with COVID-19 is asymptomatic, 0.5.

Limited data is available to give an accurate estimate on those two parameters. For simplicity, we assume *ψ* = 0.5 and let *M* vary from 0% to 1% in our global sensitivity analysis. *c*: community contact multiplier that multiplies contacts between university population and the community, 1–10.

As mentioned in Section 3.3, the numbers in ℂ^*o*^, contact with outside community, are not directly produced from known data. To assess the potential effect of contact with outside community on our modeling responses, we vary *c* from 1 to 10.

*w*: weekend multiplier that scales the increase in social interaction during weekends, 1–10.

As described in Section 3.4, we multiply the unscheduled social contact-hours we calculated for a typical weekday by *w* to simulate the increased social interaction over the weekend, such as going to a larger gathering. We vary this parameter from 1 to 10.

*p*: social percentage, 50%–150%.

Since there is a large uncertainty in the number of social contact-hours we use in ℂ^*s*^, we multiply this matrix by *p* and let it to vary from 50% to 150%.

1 − *m*: reduction in *β* with mask usage, 0%–60%.

Reduction in transmission rate of COVID-19 using masks often depends on the material and fit of the mask. [52] demonstrated mask efficiencies of approximately 20%-70%. We note these experiments were performed on mannequins and the masks were fitted correctly. To account for reality, we have reduced these rates slightly. [53] provides a review of estimates of mask efficacy for reduction in transmission for various coronaviruses (e.g., SARS) as well as modeling estimates for reduction in transmission rates for COVID-19, which demonstrated wide variations in efficacy.

*α*: probability that a symptomatic individual will self-isolate, 0%–100%. We let *α* vary from 0% to 100%, noncompliance to perfect compliance.

1/*γ*^*s*^: average duration in symptomatic infectious state before deciding whether to isolate, 2 days.

We assume that individuals make the decision to isolate when symptoms appear or the day after symptoms appear. A meta-analysis found that individuals begin being infectious a day or two before symptom onset [46], so we set 1/*γ*^*s*^ = 2.

1/*γ*^*a*^: average duration in asymptomatic infectious state, 14 days.

1/*h*: average duration of symptomatic individuals in self-isolation, 12 days.

1/*δ*: average duration of symptomatic individuals not self-isolating, 12 days.

A retrospective cohort study [54] reported that the time to recovery from symptom onset was significantly shorter in younger adults, for example the estimated median was 22.4 days (95% CI 20.8-24.1%) for individuals aged 50-59 years and 19.2 days (95% CI 17.5-21.0%) for individuals aged 20-29 years. The model-based analysis [55] estimated a mean duration from symptoms onset to hospital discharge of 24.7 days (95% credible interval 22.9-28.1). A retrospective cohort study of adult patients in Wuhan, China [56] estimated a median duration of viral shedding of 20 days (IQR 17-24) in survivors. But data obtained through hospital surveillance are likely to represent patients with moderate or severe illness. Data for outpatients with mild illness or no symptoms is needed to fully characterize the infectious period for the general population. In a multistate telephone survey of symptomatic adults conducted in the United States, 65% (*n* = 175) reported that they had returned to their usual state of health a median of 7 days (IQR 5-12) from the date of testing and among respondents aged 18-34 years with no chronic medical condition, 19% reported not having returned to their usual state of health at the time of interview [57]. [58] evaluated the clinical course of asymptomatic and mildly symptomatic patients admitted to community treatment centers (CTCs) for isolation in South Korea (average age of patients ≈ 40 years) and found that the virologic remission period was longer in symptomatic patients, 21.8 *±* 7.6 (mean *±* standard deviation) days, than in asymptomatic patients, 19.1 *±* 7.5 days; the mean number of days from symptom onset to virologic remission for patients who developed symptoms during their illness was 11.7 *±* 8.2 days. Since we consider a population that is younger and healthier than the general population, we assume a shorter period in our study, i.e., 1/*h* = 1/*δ* = 12 and 1/*γ*^*a*^ = 14 = 1/*γ*^*s*^ + 1/*h* = 1/*γ*^*s*^ + 1/*δ*. It will be interesting to see the effect of varying these parameters in a future study.

*v*_*u*,0_, *v*_*d*,0_: Percentage of *u* or *d* vaccinated at time *t* = 0, 0%–80%.

*v*_*g*,0_, *v*_*f*,0_: Percentage of *g* or *f* vaccinated at time *t* = 0, 0%–100%.

We would like to understand how vaccine hesitancy within the campus population will impact disease propagation on campus, so we vary those parameters in a wide range.

*n*_*d*_: number of on-campus undergraduates, 2885.

*n*_*u*_: number of off-campus undergraduates, 5441.

*n*_*g*_: number of graduates, 721.

*n*_*f*_ : number of faculty/staff, 424.

The faculty, student, and staff counts come directly from excel spreadsheets generated by registrar and employment data.

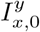: number of *y* (*s* = symptomatic, *a* = asymptomatic) infectious individuals from population *x* (*u* = off-campus undergrads, *d* = on-campus undergrads, *g* = graduate students, *f* = faculty/staff) at time *t* = 0, (0–0.5%)×*n*_*x*_.

We assume that it is very unlikely for large numbers of infectious individuals to come to campus, as many campuses require a negative test prior to allowing people to come back to campus at the beginning of the semester. To account for the false negativity of COVID-19 tests and possible reinfection since the negative test result, we let the number of symptomatic (asymptomatic) infectious individuals from population *x*, 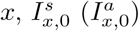, vary between 0% and 0.5% of the subpopulation size *n*_*x*_, i.e., 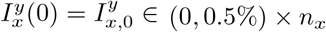 with *y* ∈ {*a, s*} and *x* ∈ {*u, d, g, f*}.

## B University Network Visualization

**Figure 11:**
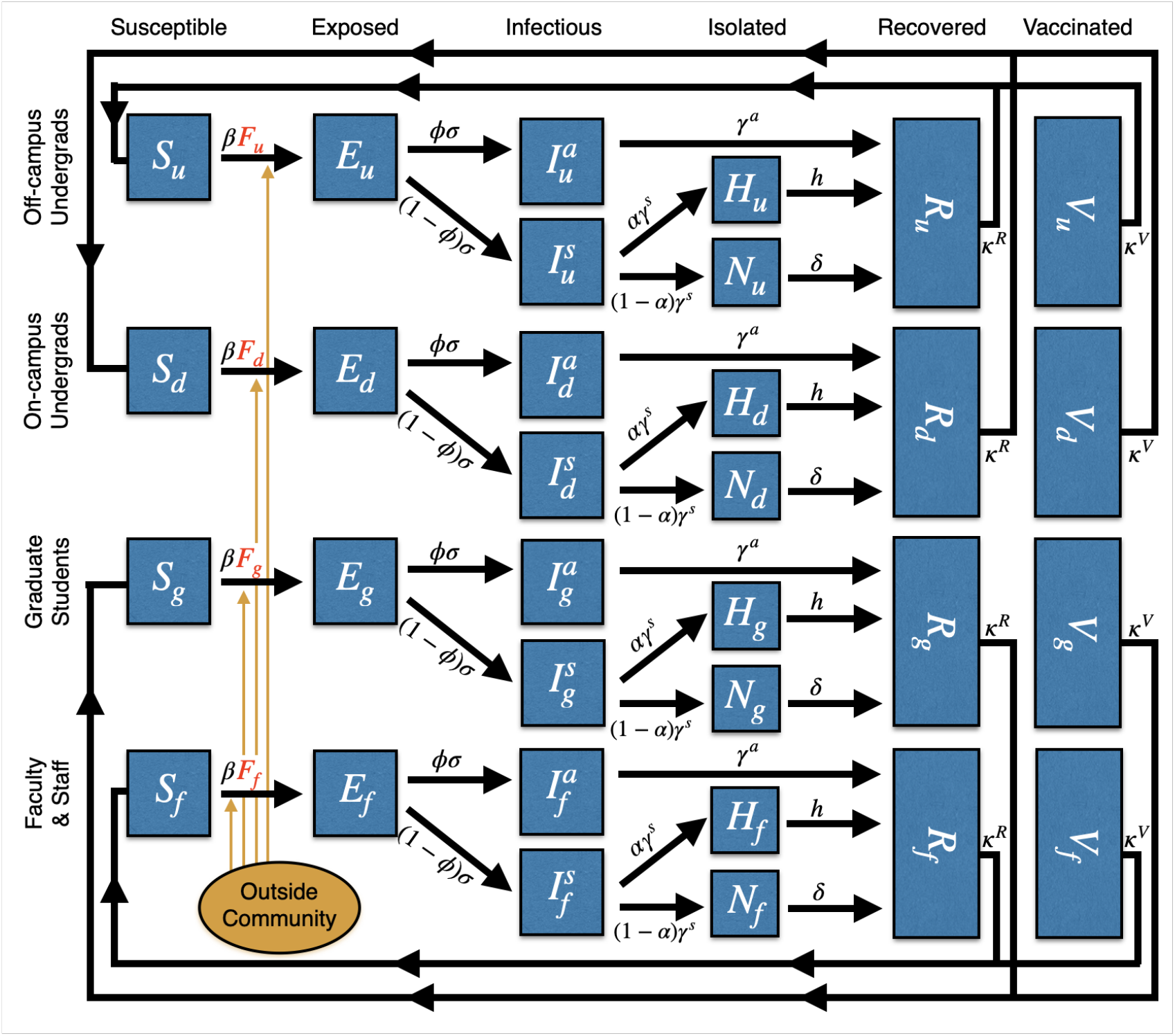
All subpopulations included in the ODE model, (1) undergraduate students living off campus (*u*), (2) undergraduate students living on-campus and in dorms (*d*), (3) graduate students (*g*), and (4) faculty and staff (*f*) and the phases of the disease that each individual progresses through: susceptible (*S*), exposed (*E*), asymptomatically infectious (*I*_*a*_) or symptomatically infectious (*I*_*s*_), if symptomatically infectious individuals can choose to self-isolate (at home) (*H*) or not (*N*), and finally both asymptomatically or symptomatically infectious recover (*R*). Note that some percentage of the population is initially vaccinated (*V*) and, for simplicity, we assume no more individuals will get vaccinated once the semester begins. Both vaccinated individuals and recovered individuals can become susceptible after certain period of time

**Figure 12:**
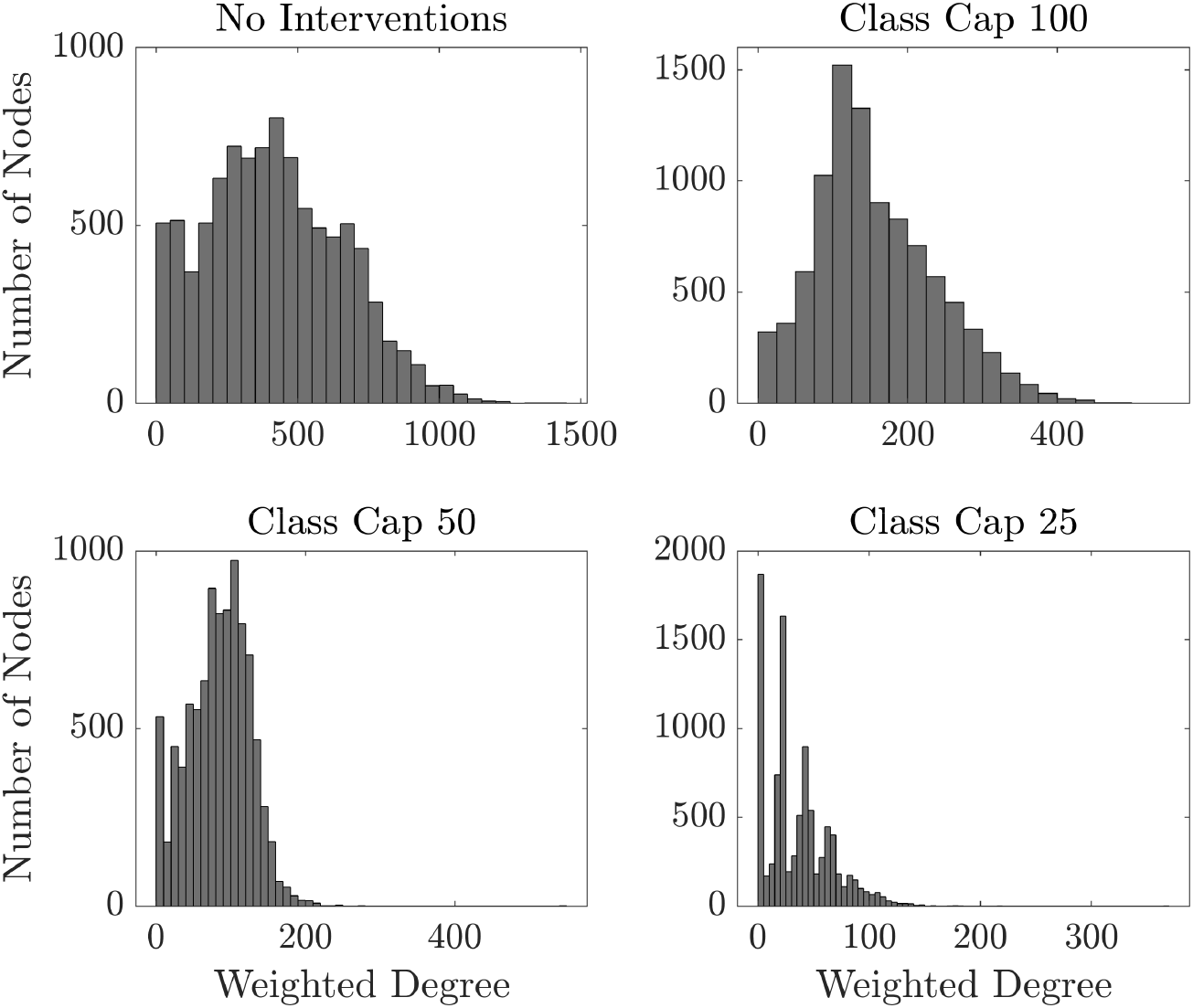
Histogram of weighted degrees for the university network under four interventions with different class cap sizes. The upper left panel contains no interventions and the lower right panel assumes classes that are larger than 25 students do not meet in person. Bin width are 50, 25, 10 and 5 for No Intervention, class cap 100, class cap 50, and class cap 25, respectively

**Figure 13:**
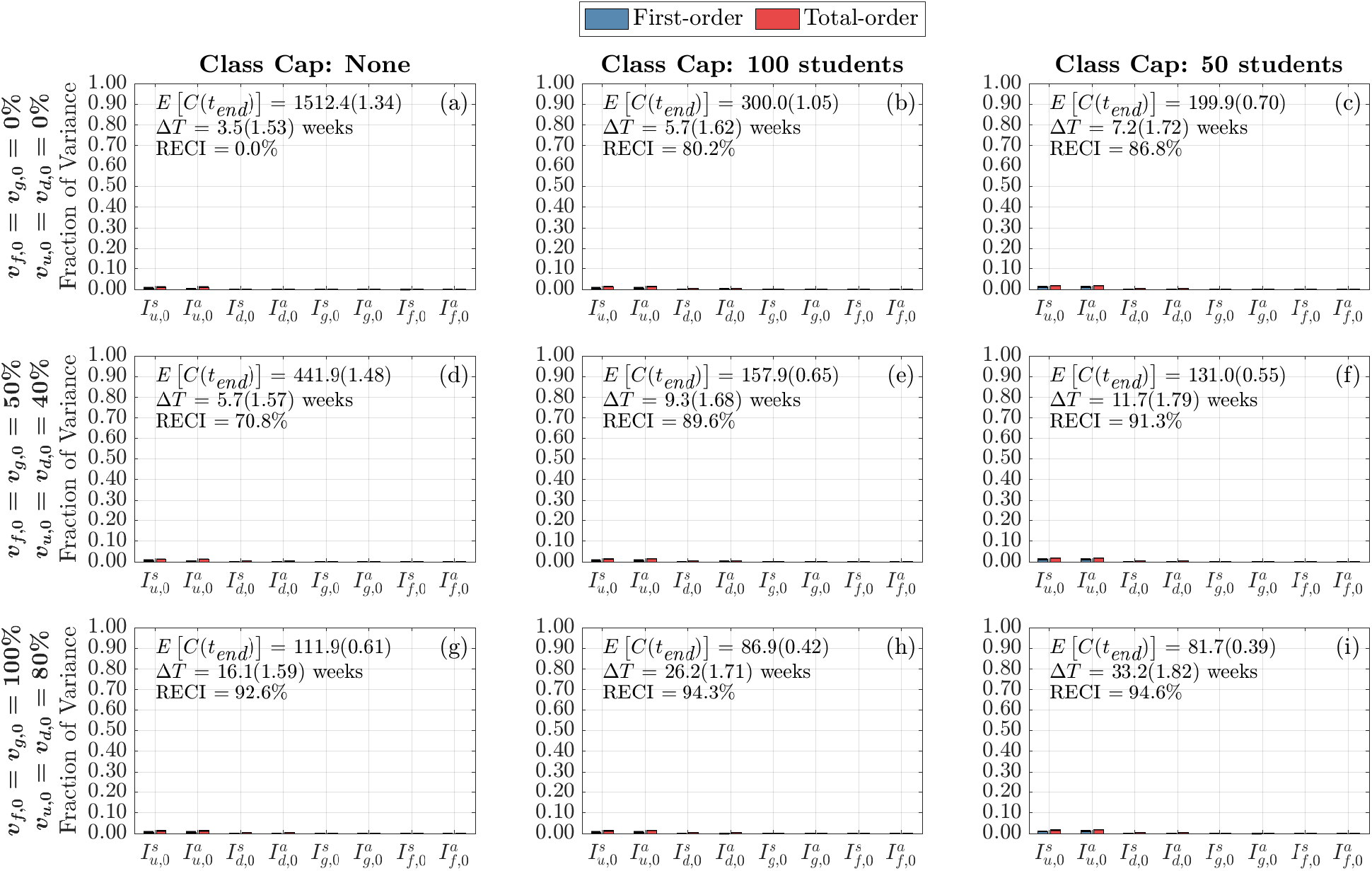
Global Sensitivity Analysis of Doubling Time to Initial Conditions. First-order (blue) and total-order (red) are shown as well as the standard errors. The mean and coefficient of variation of the doubling time and total cumulative infections are reported in each subplot. Each column represents three class cap scenarios: none, 100 student, and 50 student caps. Each row represents one of three vaccination scenarios at the start of the semester. First row: 0% vaccination; second row: 50% of faculty, 50% graduate students, and 40% of undergraduate students vaccinated; third row: 100% of faculty, 100% graduate students, and 80% of undergraduate students vaccinated

**Figure 14:**
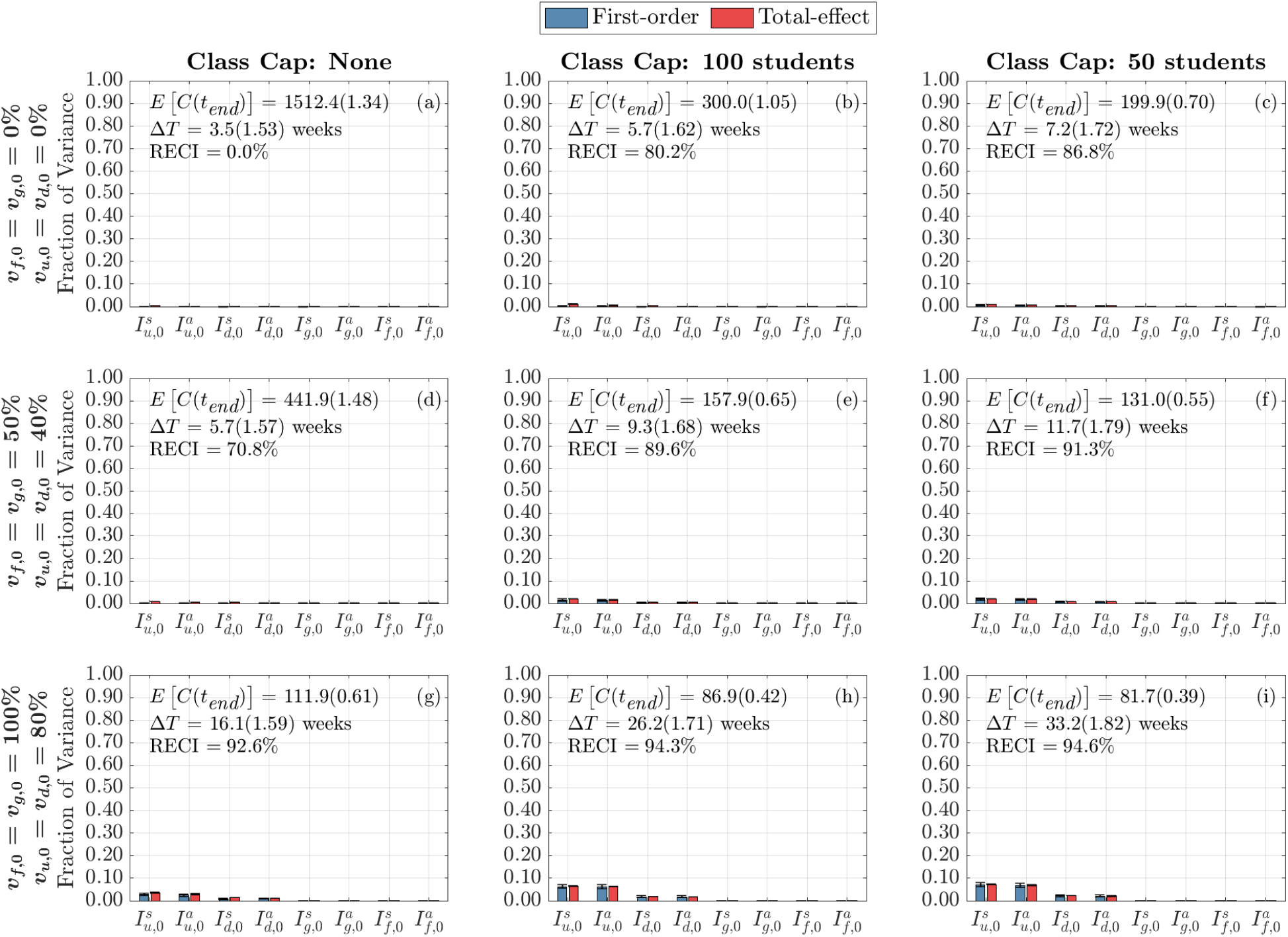
Global Sensitivity Analysis of Initial Conditions on Cumulative Infections at the End of the Term. Each column represents three class cap scenarios: none, 100 student, and 50 student caps. Each row represents one of three vaccination scenarios at the start of the semester. First row: 0% vaccination; second row: 50% of faculty, 50% graduate students, and 40% of undergraduate students vaccinated; third row: 100% of faculty, 100% graduate students, and 80% of undergraduate students vaccinated

**Figure 15:**
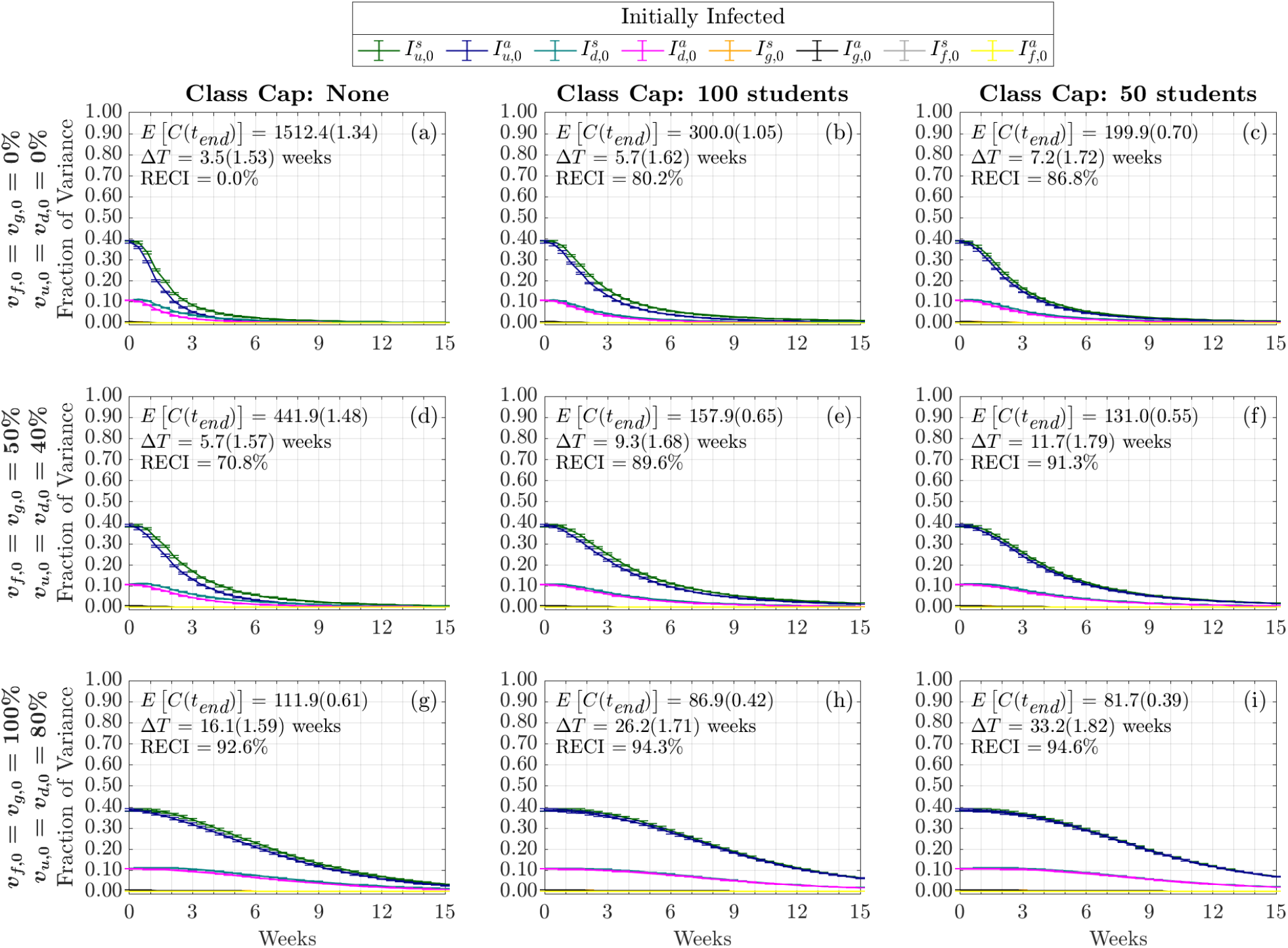
Time-Varying Total-Order Effect of Initial Conditions on Cumulative Infections. The total-order Sobol indices are shown as well as the standard errors. The mean and coefficient of variation for the doubling time and total cumulative infections are reported in each subplot. Each column represents three class cap scenarios: none, 100 student, and 50 student caps. Each row represents one of three vaccination scenarios at the start of the semester. First row: 0% vaccination; second row: 50% of faculty, 50% graduate students, and 40% of undergraduate students vaccinated; third row: 100% of faculty, 100% graduate students, and 80% of undergraduate students vaccinated

**Figure 16:**
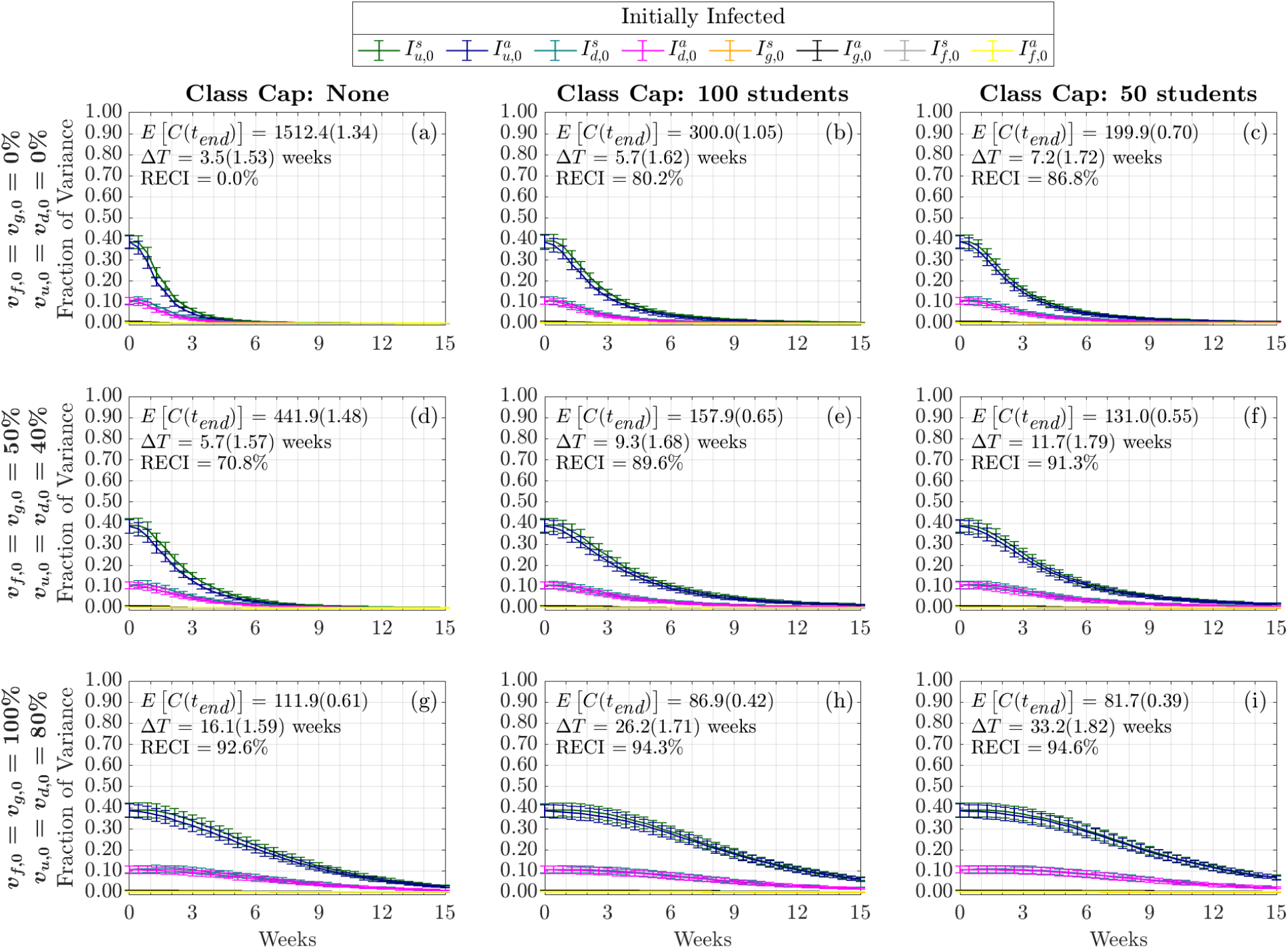
Time-Varying First-Order Effect of Initial Conditions on Cumulative Infections. Each column represents three class cap scenarios: none, 100 student, and 50 student caps. Each row represents one of three vaccination scenarios at the start of the semester. First row: 0% vaccination; second row: 50% of faculty, 50% graduate students, and 40% of undergraduate students vaccinated; third row: 100% of faculty, 100% graduate students, and 80% of undergraduate students vaccinated

## C Figures of Sensitivity Analysis to Initial Conditions

## D Details of the Contact Matrix

Here we discuss in more detail the calculation of the contact matrix ℂ. With examples for our baseline case (no class cap), we explain the calculation of the classroom contact matrix ℂ^*c*^, the living contact matrix ℂ^*l*^, the contact matrix for outside community and campus subpopulations ℂ^*o*^, and the social contact matrix ℂ^*s*^.

Table 5 displays the classroom contact matrix created using the registrar data with no class caps, ℂ^*c*^. Note that since the outside community does not engage in courses, the entries for the fifth column of this sub-matrix are 0. Except for faculty & staff, most other subpopulations have contact-hours that are higher within their own subpopulation. This intuitively makes sense since most students take courses with those in their subpopulation. In general, lower-level courses are taken by students who are in the freshman or sophomore stage, who also live on-campus. Upper-level courses tend to be taken by students more advanced in their college degree, who tend to live off-campus.

**Table 5:**
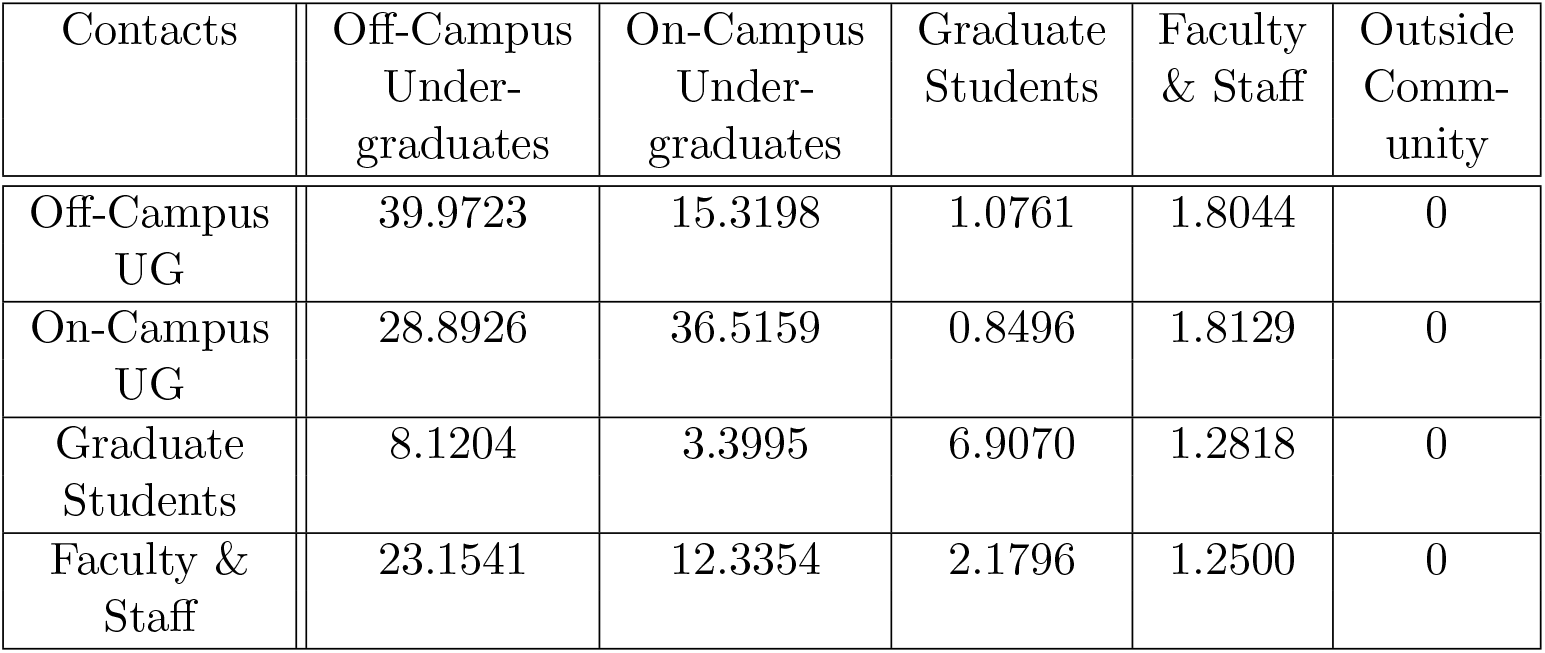
The contact matrix for in-class instruction, ℂ^*c*^, calculated using registrar data between the subpopulations off-campus undergraduates, on-campus undergraduates, graduate students, and faculty & staff under no class caps, assuming far contacts are 25% less likely to transmit than close contacts.

One may notice that these classroom contact-hours do not appear to be symmetric. We note that, in the model, contact-hours are scaled by the size of the subpopulation (*n*_*u*_ = 5441, *n*_*d*_ = 2885, *n*_*g*_ = 721, *n*_*f*_ = 424). When examining the various subpopulations, we can see that the contact-hours are symmetric between those populations (e.g., 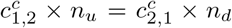). This symmetry ensures that the contact-hours are reciprocal.

We can also examine the sub-contact matrix generated by living situations (living in dorms or apartments), ℂ^*l*^. As shown in Table 6, the first four columns of this matrix is diagonal and, thus, symmetric, as the various subpopulations only live with the same subpopulation. The details for calculating these living contact-hours are described in Section 3.2.

**Table 6:** The contact matrix for living situations, ℂ^*l*^.

| Contacts | Off-Campus Under-graduates | On-Campus Under-graduates | Graduate Students | Faculty & Staff | Outside Community |
| --- | --- | --- | --- | --- | --- |
| Off-Campus UG | 12 | 0 | 0 | 0 | 0 |
| On-Campus UG | 0 | 21.5472 | 0 | 0 | 0 |
| Graduate Students | 0 | 0 | 6 | 0 | 0 |
| Faculty & Staff | 0 | 0 | 0 | 0 | 0 |

Beyond the university, subpopulations interact with the surrounding community. Table 7 displays the contacts between each of the subpopulations and the outside community. Since we do not directly model the outside community, these interactions occur solely in the fifth column of the contact matrix. See Section 3.3 for more information regarding the choices of values.

**Table 7:** The contact matrix for contacts between campus subpopulations and outside community, ℂ^*o*^.

| Contacts | Off-Campus Under-graduates | On-Campus Under-graduates | Graduate Students | Faculty & Staff | Outside Community |
| --- | --- | --- | --- | --- | --- |
| Off-Campus UG | 0 | 0 | 0 | 0 | 5 |
| On-Campus UG | 0 | 0 | 0 | 0 | 1 |
| Graduate Students | 0 | 0 | 0 | 0 | 5 |
| Faculty & Staff | 0 | 0 | 0 | 0 | 15 |

In addition to the matrices for in-class instruction, living situation, and outside community, there is also the ‘social’ matrix ℂ^*s*^. In this case, it is assumed that only undergraduate students engage in unplanned social gatherings. To construct our social sub-matrix, we multiply the classroom contact-hours in Table 5 by 0.3, as described in Section 3.4. Table 8 displays our nominal calculation for the social sub-matrix for a typical weekday. Recall, as explained in Section 3.4, during the weekend, we multiply this social matrix by a multiplier, *w*, to represent larger weekend gatherings such as parties. As with the classroom contacts, the social contacts are reciprocal between subpopulations.

**Table 8:** The contact matrix for unplanned social contacts (e.g., parties or study groups), ℂ^*s*^, on a typical weekday.

| Contacts | Off-Campus Under-graduates | On-Campus Under-graduates | Graduate Students | Faculty & Staff | Outside Community |
| --- | --- | --- | --- | --- | --- |
| Off-Campus UG | 11.9917 | 4.5960 | 0 | 0 | 0 |
| On-Campus UG | 8.6678 | 10.9548 | 0 | 0 | 0 |
| Graduate Students | 0 | 0 | 0 | 0 | 0 |
| Faculty & Staff | 0 | 0 | 0 | 0 | 0 |

Next we present the full contact matrix, C, which is sum of all the submatrices shown above (i.e., ℂ = ℂ^*c*^ + ℂ^*l*^ + ℂ^*o*^ + ℂ^*s*^). Table 9 displays the full contract matrix under nominal conditions, assuming a far contact classroom rate of 25%.

**Table 9:** The full contact matrix ℂ under nominal conditions between the subpopulations off-campus undergraduates, on-campus undergraduates, graduate students, faculty & staff, and the outside community on a typical weekday.

| Contacts | Off-Campus Under-graduates | On-Campus Under-graduates | Graduate Students | Faculty & Staff | Outside Community |
| --- | --- | --- | --- | --- | --- |
| Off-Campus UG | 81.9640 | 19.9158 | 1.0761 | 1.8044 | 5 |
| On-Campus UG | 37.5604 | 73.0387 | 0.8496 | 1.8129 | 1 |
| Graduate Students | 8.1204 | 3.3995 | 21.9070 | 1.2818 | 5 |
| Faculty & Staff | 23.1541 | 12.3354 | 2.1796 | 1.25 | 15 |

## Supplementary File 1

### A Variance and Global Sensitivity Analysis

The formulation for the spread of COVID-19 in a classroom, detailed in the main text, distinguishes between close (droplets) and far (aerosols) transmission (see Section 3.1 for details). In this document we present the variance and Sobol analysis results with a 10% chance of infection from ‘far’ contacts.

**Fig. A1.**
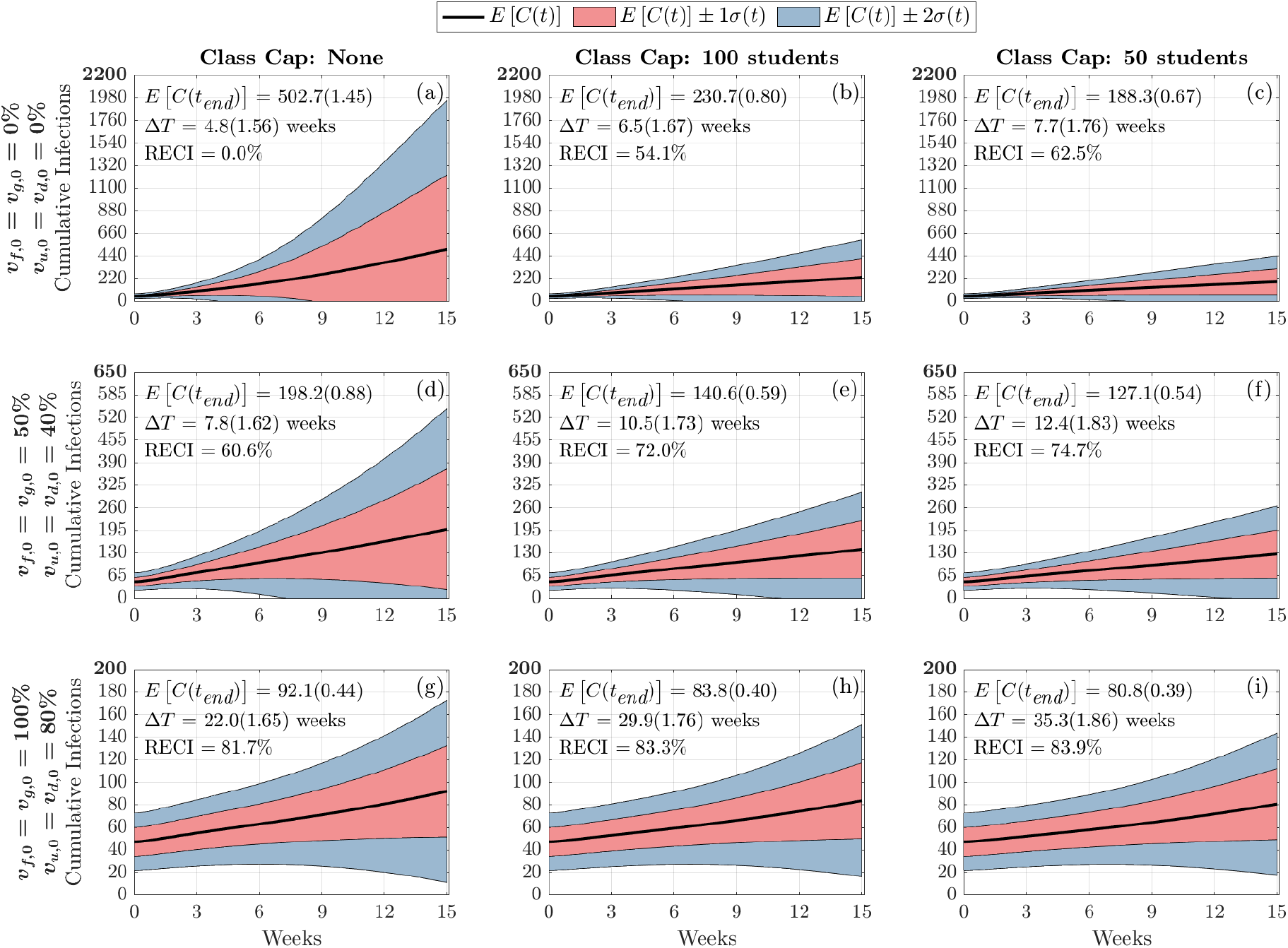
Distribution of Cumulative Infections. Figures show the distribution of cumulative infections over the span of a semester (15 weeks) and we allow the contact, infection parameters, and initial number of infectious individuals to vary (See Table 1 in main text). The mean and coefficient of variation for the doubling time and total cumulative infections are reported in each subplot. The reduction in expected cumulative infections (RECI) from case (a) with no vaccination and no class caps is presented in each subsequent case.

**Fig. A2.**
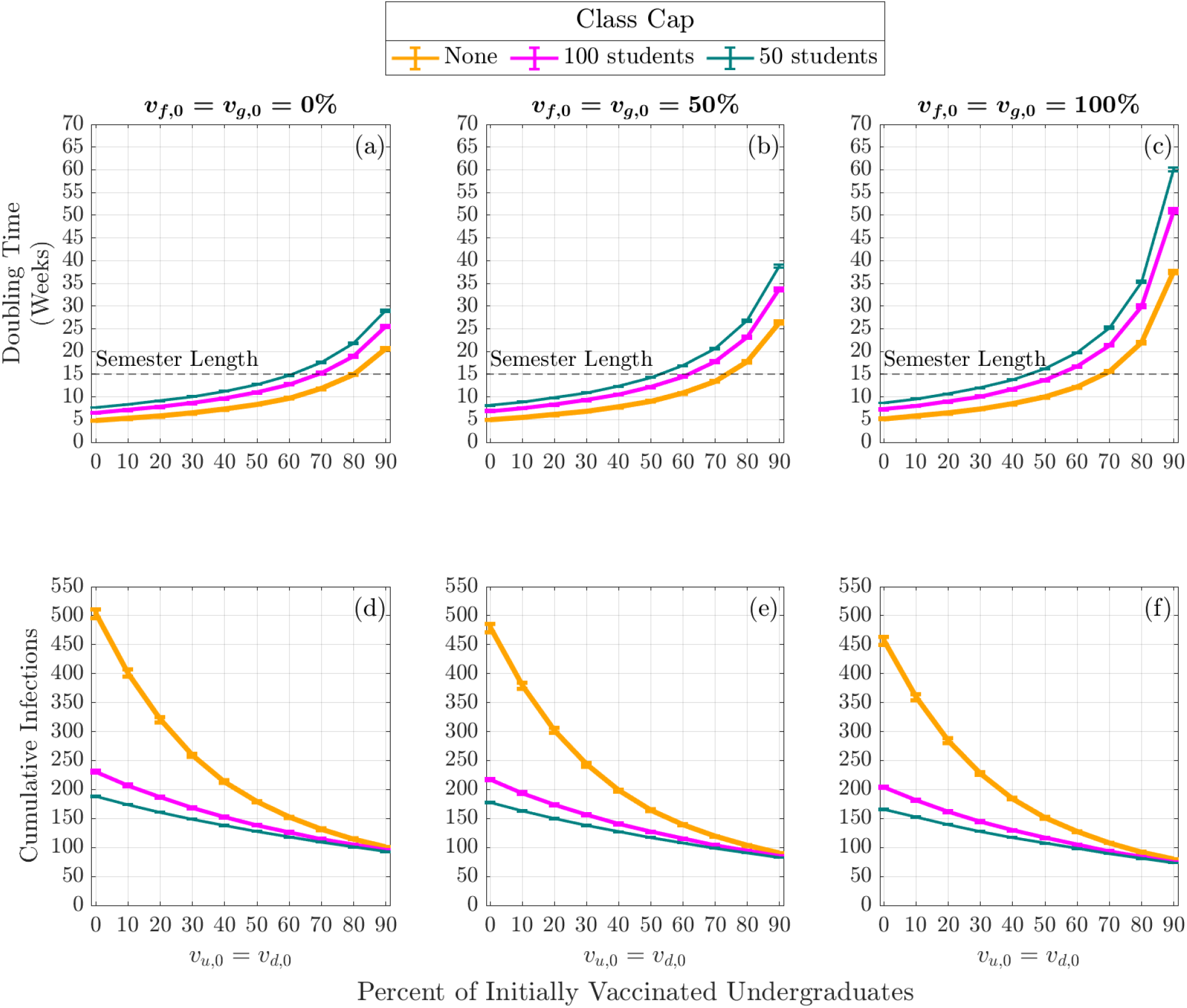
Expected Infection Doubling Times and Cumulative Infections by Class Capacity and Percent of Vaccinated Undergraduates. The expected cumulative number of infection (cumulative infections) by the end of the semester and the expected doubling time computed during the first four weeks of the semester. The error bars are a 95% confidence interval.

**Fig. A3.**
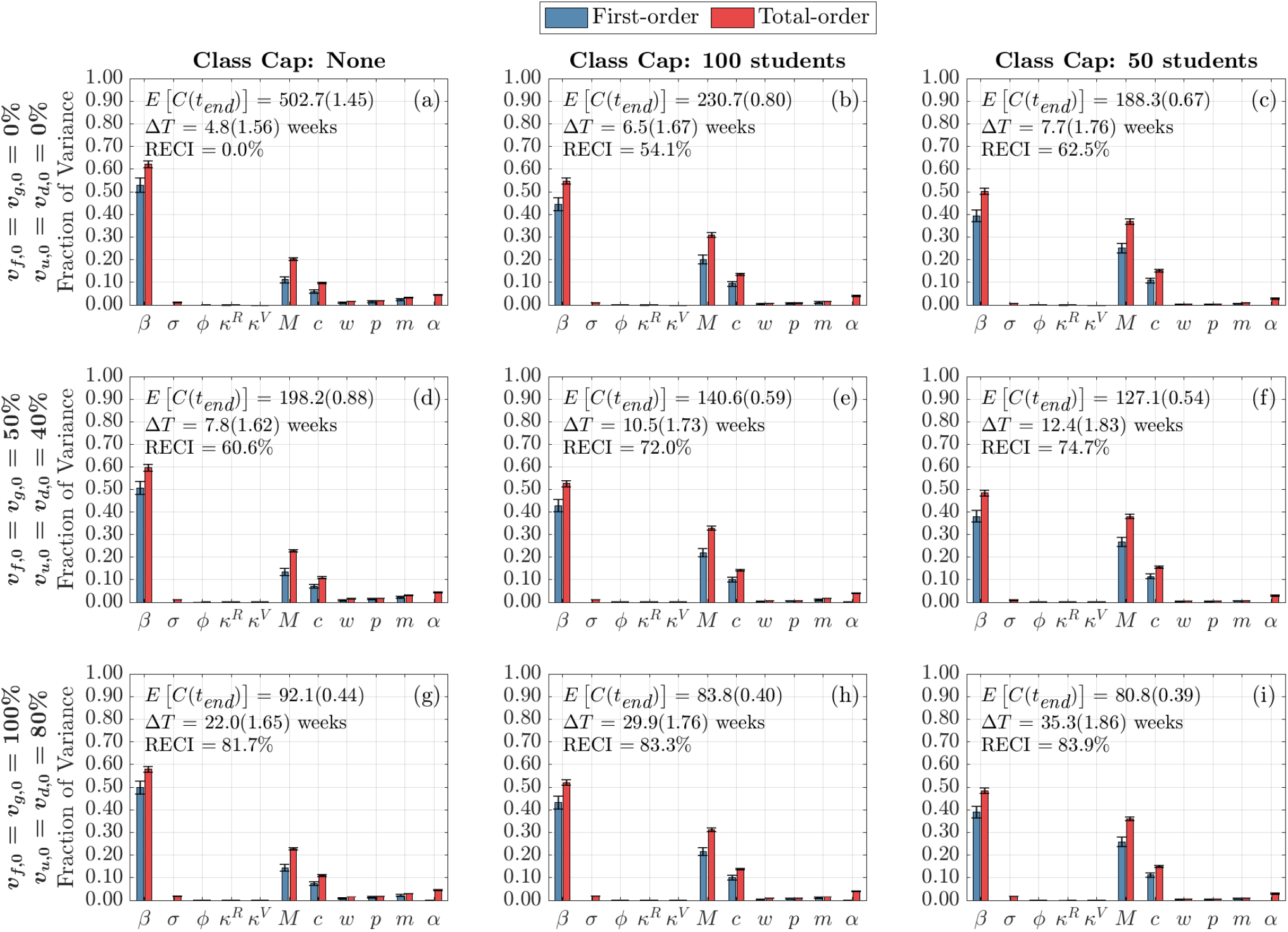
Global Sensitivity Analysis of Infection and Contact Parameters on Infection Doubling Time. First-order (blue) and total-order (red) Sobol Indices are shown as well as the standard errors. The mean and coefficient of variation for the doubling time and total cumulative infections are reported in each subplot. Each column represents three class cap scenarios: none, 100 student, and 50 student caps. Each row represents one of three vaccination scenarios at the start of the semester. First row: 0% vaccination; second row: 50% of faculty, 50% graduate students, and 40% of under-graduate students vaccinated; third row: 100% of faculty, 100% graduate students, and 80% of undergraduate students vaccinated.

**Fig. A4.**
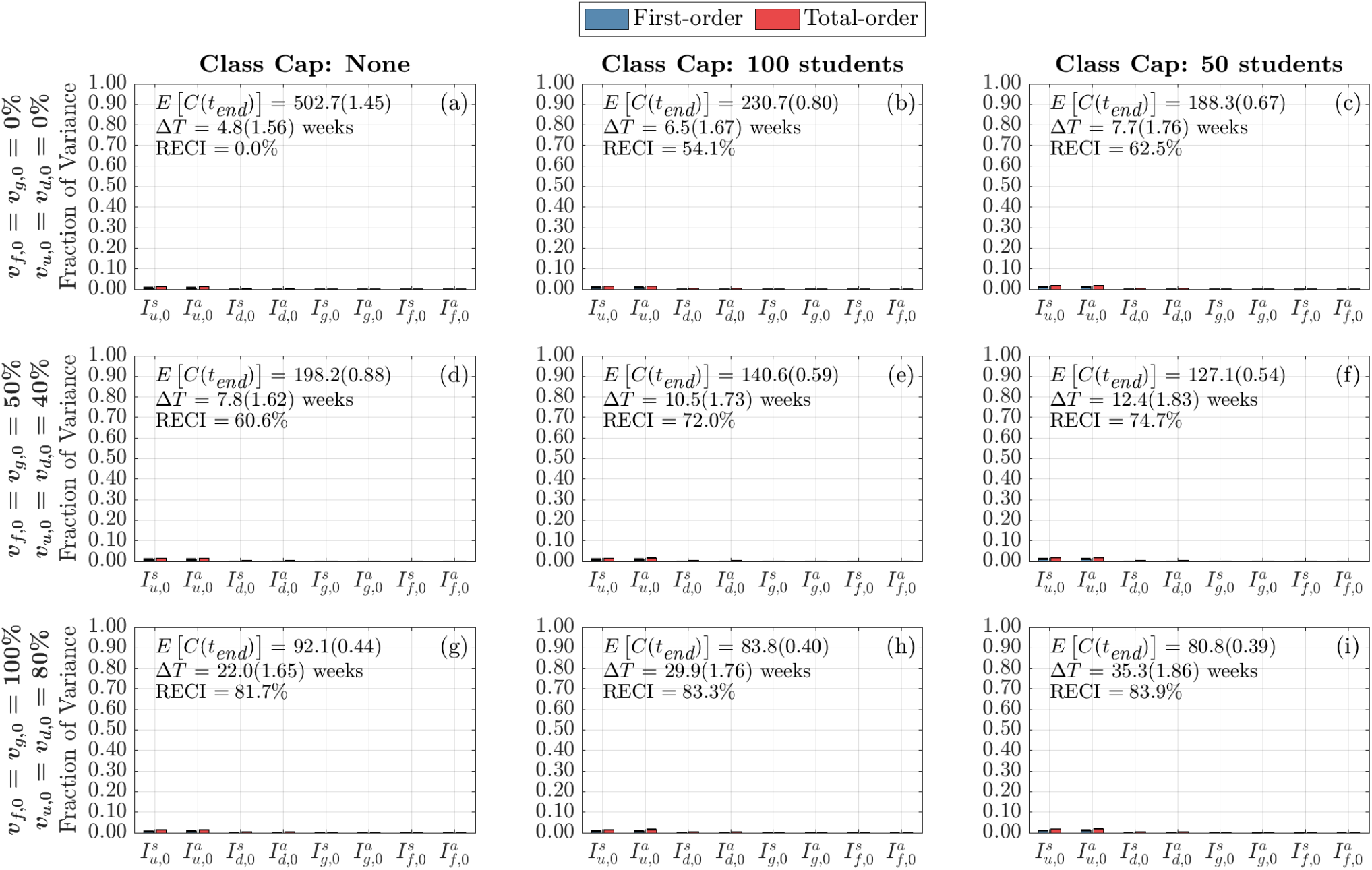
Global Sensitivity Analysis of Doubling Time to Initial Conditions. First-order (blue) and total-order (red) Sobol Indices are shown as well as the standard errors. The mean and coefficient of variation for the doubling time and total cumulative infections are reported in each subplot. Each column represents three class cap scenarios: none, 100 student, and 50 student caps. Each row represents one of three vaccination scenarios at the start of the semester. First row: 0% vaccination; second row: 50% of faculty, 50% graduate students, and 40% of under-graduate students vaccinated; third row: 100% of faculty, 100% graduate students, and 80% of undergraduate students vaccinated.

**Fig. A5.**
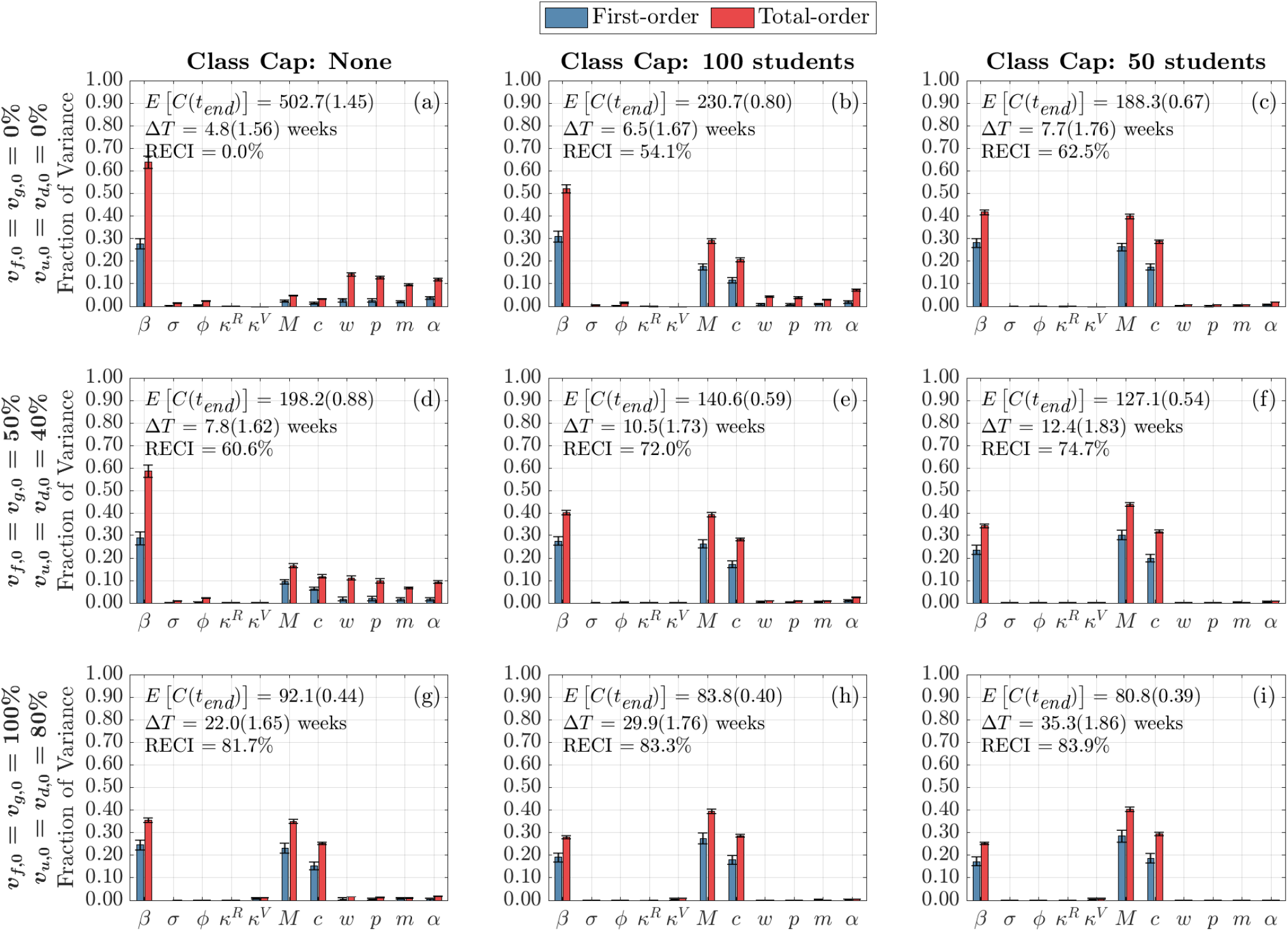
Global Sensitivity Analysis of Infection and Contact Parameters on Cumulative Infections at the End of the Term. Each column represents three class cap scenarios: none, 100 student, and 50 student caps. Each row represents one of three vaccination scenarios at the start of the semester. First row: 0% vaccination; second row: 50% of faculty, 50% graduate students, and 40% of undergraduate students vaccinated; third row: 100% of faculty, 100% graduate students, and 80% of undergraduate students vaccinated.

**Fig. A6.**
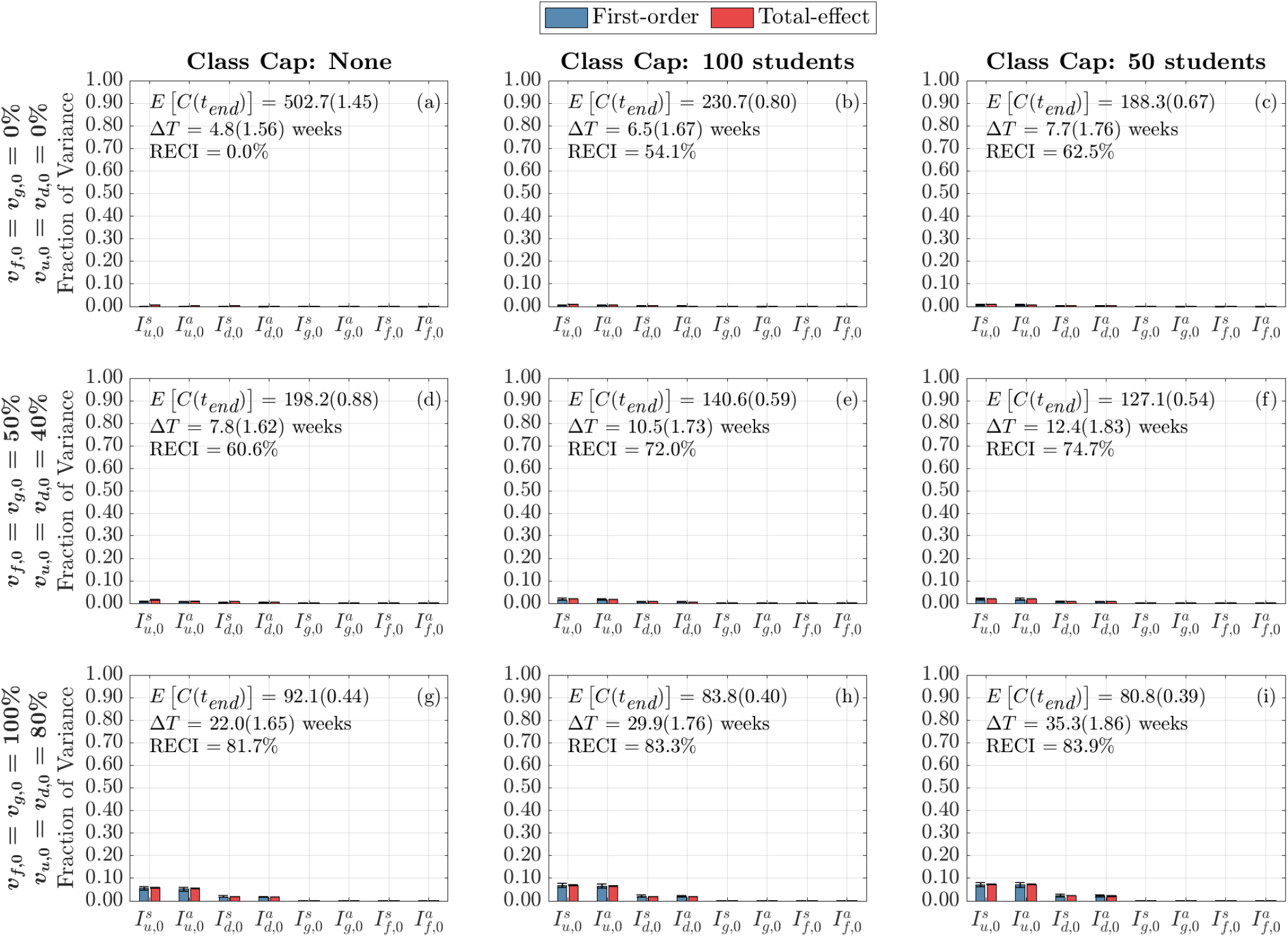
Global Sensitivity Analysis of Initial Conditions on Cumulative Infections at the End of the Term. Each column represents three class cap scenarios: none, 100 student, and 50 student caps. Each row represents one of three vaccination scenarios at the start of the semester. First row: 0% vaccination; second row: 50% of faculty, 50% graduate students, and 40% of undergraduate students vaccinated; third row: 100% of faculty, 100% graduate students, and 80% of undergraduate students vaccinated.

**Fig. A7.**
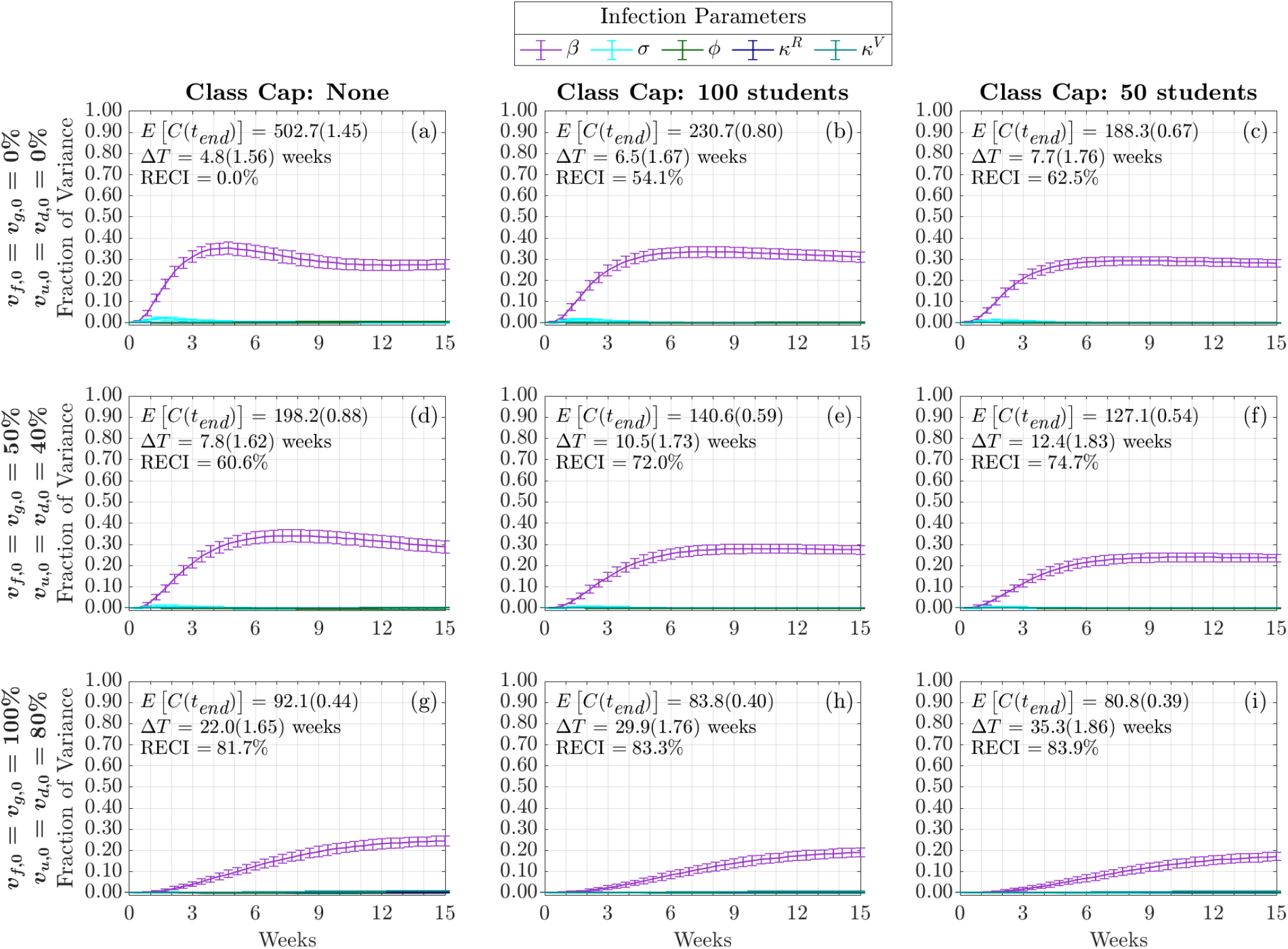
Time-Varying First-Order Effect of Infection Parameters on Cumulative Infections. Each column represents three class cap scenarios: none, 100 student, and 50 student caps. Each row represents one of three vaccination scenarios at the start of the semester. First row: 0% vaccination; second row: 50% of faculty, 50% graduate students, and 40% of undergraduate students vaccinated; third row: 100% of faculty, 100% graduate students, and 80% of undergraduate students vaccinated.

**Fig. A8.**
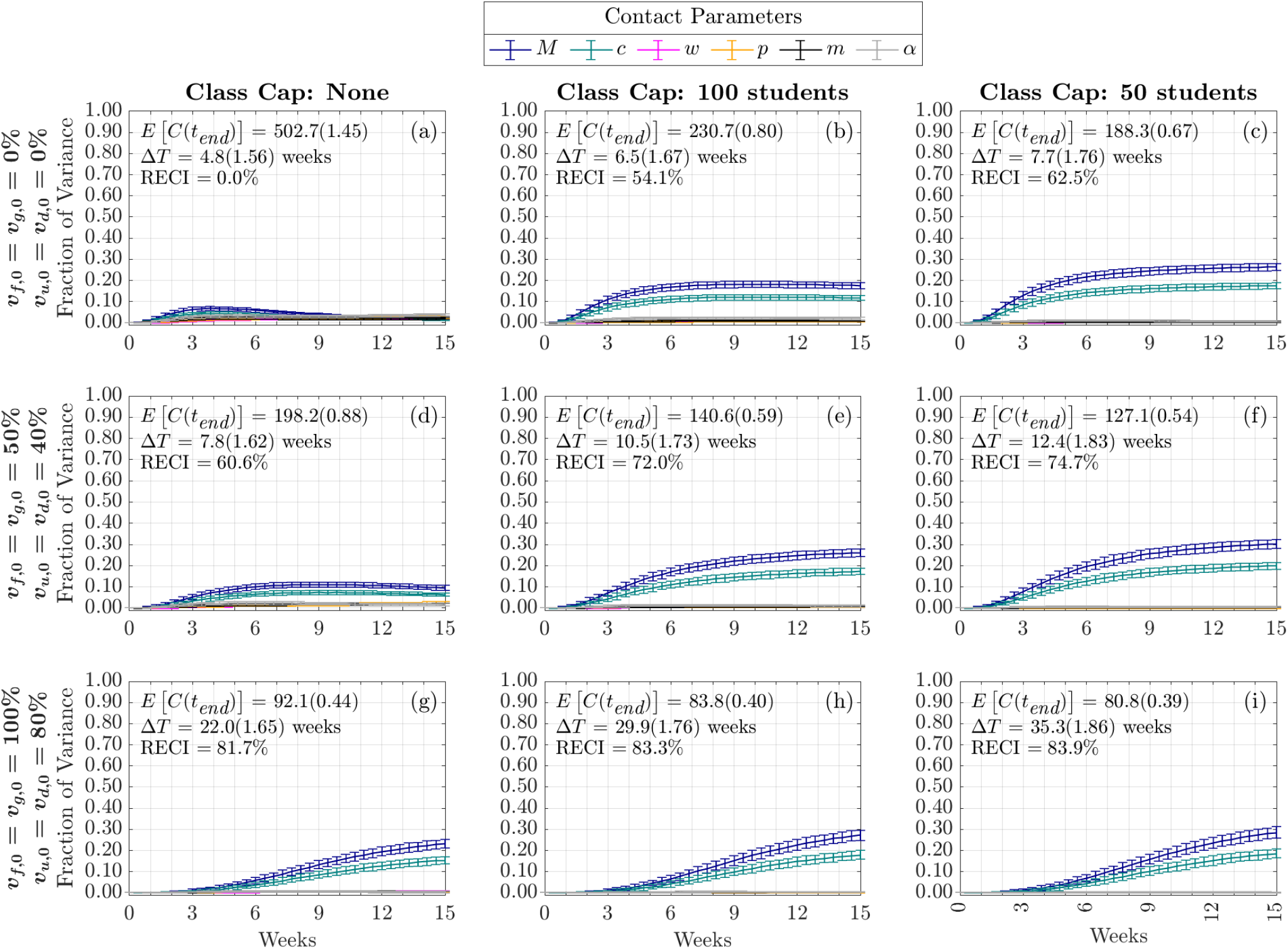
Time-Varying First-Order Effect of Contact Parameters on Cumulative Infections. Each column represents three class cap scenarios: none, 100 student, and 50 student caps. Each row represents one of three vaccination scenarios at the start of the semester. First row: 0% vaccination; second row: 50% of faculty, 50% graduate students, and 40% of undergraduate students vaccinated; third row: 100% of faculty, 100% graduate students, and 80% of undergraduate students vaccinated.

**Fig. A9.**
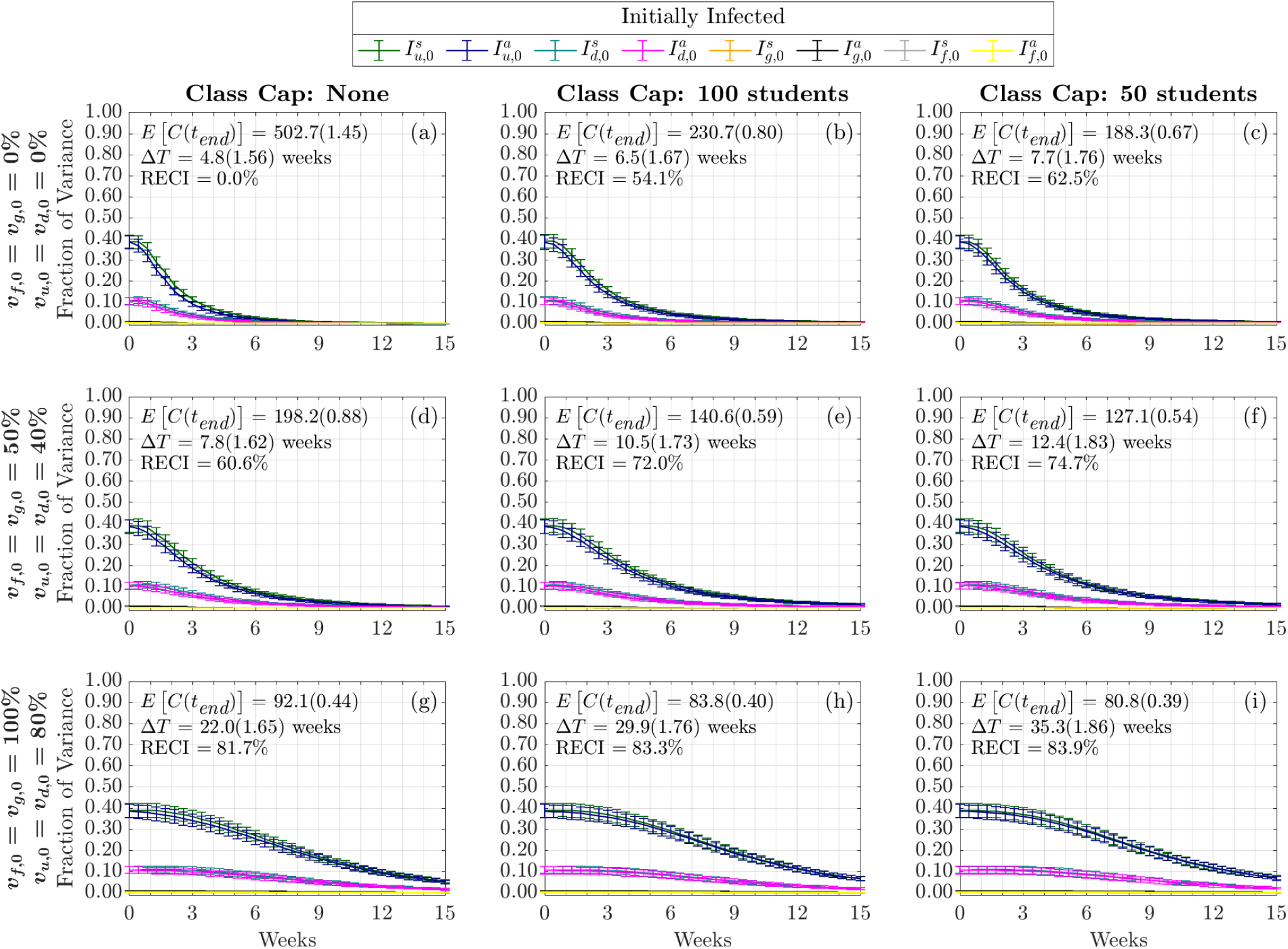
Time-Varying First-Order Effect of Initial Conditions on Cumulative Infections. Each column represents three class cap scenarios: none, 100 student, and 50 student caps. Each row represents one of three vaccination scenarios at the start of the semester. First row: 0% vaccination; second row: 50% of faculty, 50% graduate students, and 40% of undergraduate students vaccinated; third row: 100% of faculty, 100% graduate students, and 80% of undergraduate students vaccinated.

**Fig. A10.**
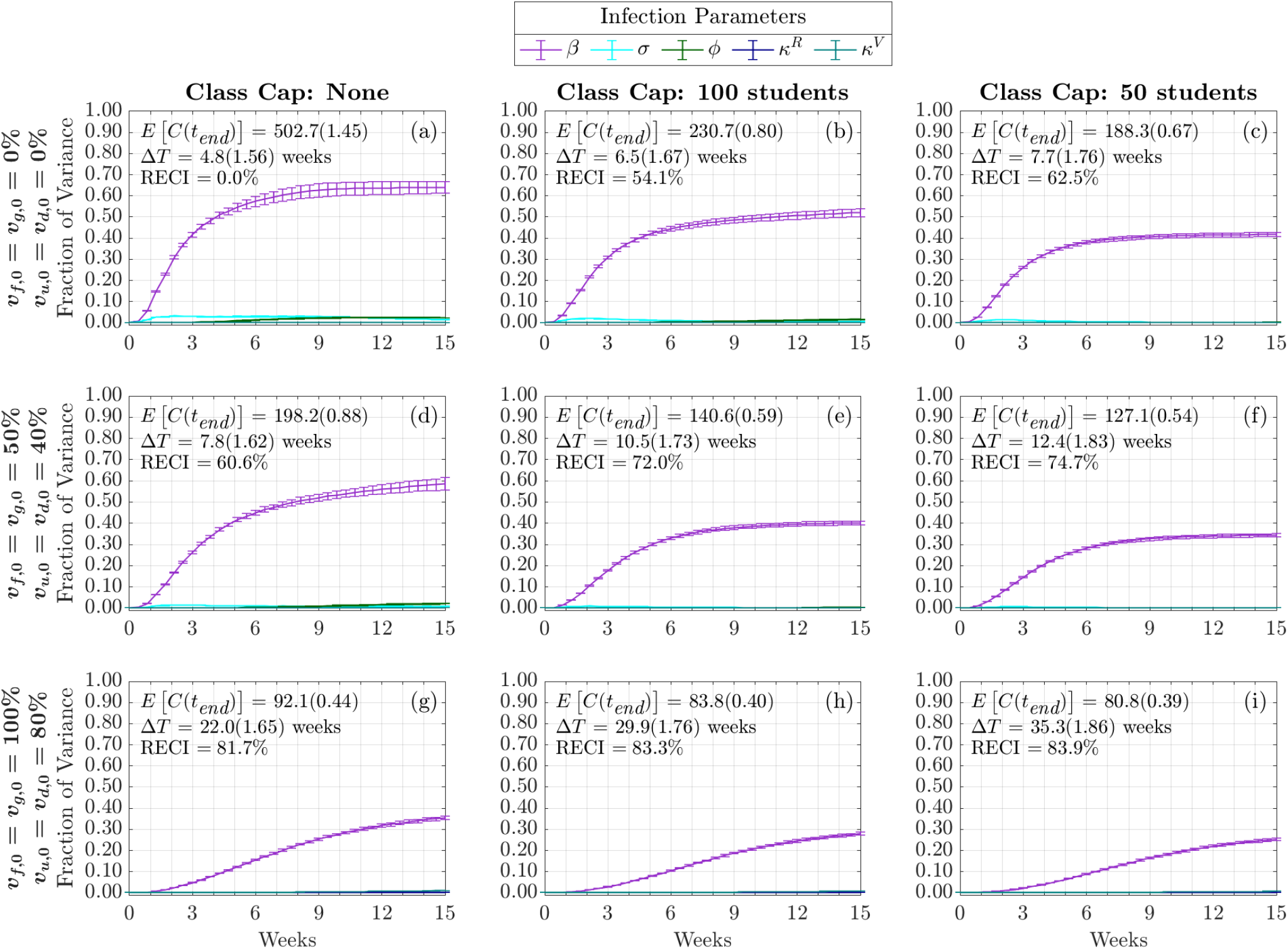
Time-Varying Total-Order Effect of Infection Parameters on Cumulative Infections. Each column represents three class cap scenarios: none, 100 student, and 50 student caps. Each row represents one of three vaccination scenarios at the start of the semester. First row: 0% vaccination; second row: 50% of faculty, 50% graduate students, and 40% of undergraduate students vaccinated; third row: 100% of faculty, 100% graduate students, and 80% of undergraduate students vaccinated.

**Fig. A11.**
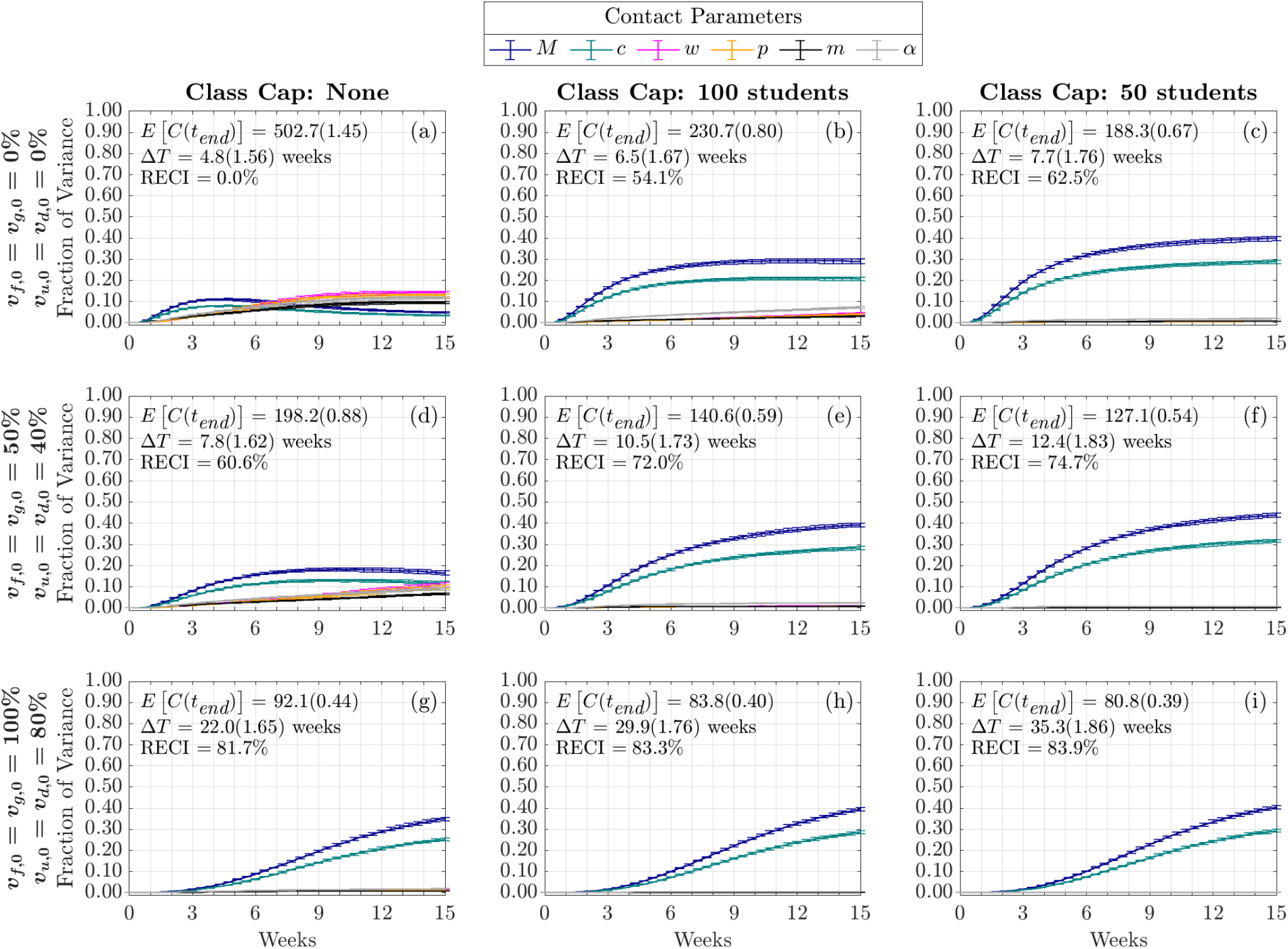
Time-Varying Total-Order Effect of Contact Parameters on Cumulative Infections. Each column represents three class cap scenarios: none, 100 student, and 50 student caps. Each row represents one of three vaccination scenarios at the start of the semester. First row: 0% vaccination; second row: 50% of faculty, 50% graduate students, and 40% of undergraduate students vaccinated; third row: 100% of faculty, 100% graduate students, and 80% of undergraduate students vaccinated.

**Fig. A12.**
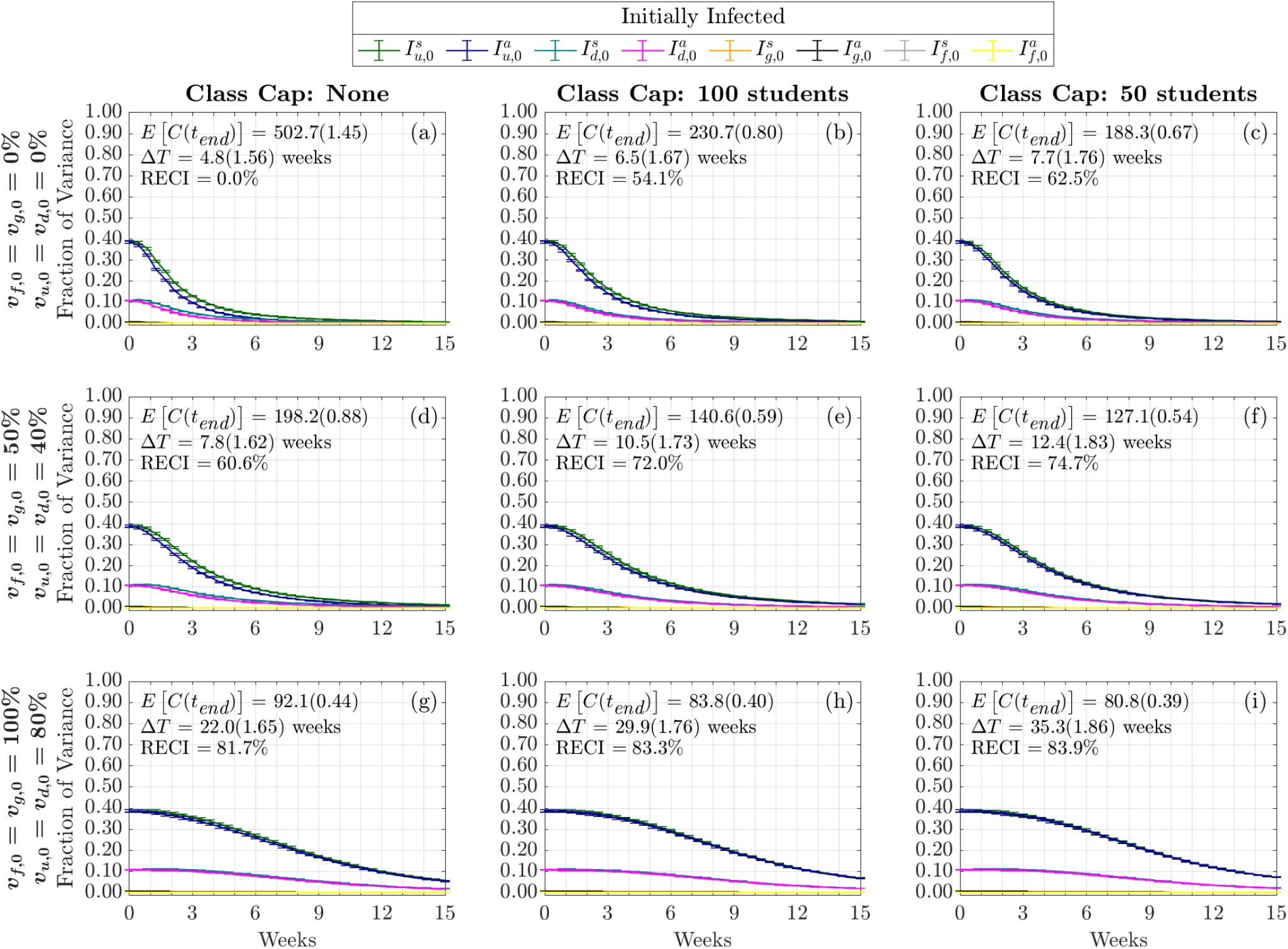
Time-Varying First-Order Effect of Initial Conditions on Cumulative Infections. Each column represents three class cap scenarios: none, 100 student, and 50 student caps. Each row represents one of three vaccination scenarios at the start of the semester. First row: 0% vaccination; second row: 50% of faculty, 50% graduate students, and 40% of undergraduate students vaccinated; third row: 100% of faculty, 100% graduate students, and 80% of undergraduate students vaccinated.

## Supplementary File 2

### B Network Visualization

Visualization of the university network as a weighted undirected network under four interventions with different class cap size. Edge thickness reflects weights (minimum 1 and maximum 10) **where each edge represents a classroom contact. In these figures, only classroom contacts are shown**. Nodes were colored according to their roles (reddish purple for off-campus undergraduate students, yellow for on-campus undergraduate students, sky blue for graduate students, and vermillion for faculty/staff) and sized according to weighted degree (smaller nodes correspond to smaller weighted degrees). See Table 4 for the breakdown of the networks. Those four figures were all generated using the same random seed under the same settings. Note that those four figures are provided as four individual Supplementary Files in PDF format (via clickable links below) to allow the interested reader to zoom in and inspect different regions of the network. It may take a while for your PDF viewer to load due to their size.

**Table 4:** University network characteristics under four interventions with different class cap size. For all four cases, the total number of nodes is 9471, with *n*_*u*_ = 5441, *n*_*d*_ = 2885, *n*_*g*_ = 721, and *n*_*f*_ = 424.

|  | No Intervention | Class Cap 100 | Class Cap 50 | Class Cap 25 |
| --- | --- | --- | --- | --- |
| Total # of isolated nodes | 0 | 10 | 305 | 1626 |
| Isolated $u$ | 0 | 2 | 253 | 1324 |
| Isolated $d$ | 0 | 0 | 18 | 222 |
| Isolated $g$ | 0 | 0 | 1 | 10 |
| Isolated $f$ | 0 | 8 | 33 | 70 |
| Weighted Degree | [1, 1417] | [0, 543] | [0, 543] | [0, 367] |
| Average Weighted Degree | 417.552 | 155.553 | 83.150 | 34.840 |
| # of unique edges | 1613685 | 614622 | 343204 | 138890 |
| Edge weights | [1, 10] | [1, 9] | [1, 8] | [1, 7] |

FigureB1 NetworkVisual NoClassCap.pdf

Visualization of university network as a weighted undirected network with no class caps, i.e., all classes meet in-person regardless of the number of registered students.

Clickable link to the figure

- FigureB2 NetworkVisual ClassCap100.pdf

Visualization of university network as a weighted undirected network with class cap 100, i.e., all classes with no more than 100 students registered meet in-person.

Clickable link to the figure

- FigureB3 NetworkVisual ClassCap50.pdf

Visualization of university network as a weighted undirected network with class cap 50, i.e., all classes with no more than 50 students registered meet in-person.

Clickable link to the figure

- FigureB4 NetworkVisual ClassCap25.pdf

Visualization of university network as a weighted undirected network with class cap 25, i.e., all classes with no more than 25 students registered meet in-person.

Clickable link to the figure

## References

[1] S. Zheng-Li, “Origins of SARS-CoV-2: focusing on science,” Infectious Diseases & Immunity, vol. 1, no. 01, pp. 3–4, 2021.

[2] World Health Organization, WHO Director-General’s opening remarks at themedia briefing on COVID-19, March 11, 2020 (accessed June 28, 2020). https://www.who.int/dg/speeches/detail/who-director-general-s-opening-remarks-at-the-media-briefing-on-covid-19-11-march-2020.

[3] Johns Hopkins Corona Virus Resource Center, COVID-19 Dashboard, 2020 (accessed October 13, 2022). https://coronavirus.jhu.edu/map.html.

[4] E. Mathieu, H. Ritchie, E. Ortiz-Ospina, M. Roser, J. Hasell, C. Appel, C. Giattino, and L. Rodés-Guirao, “A global database of COVID-19 vaccinations,” Nat. Hum. Behav., pp. 1–7, 2021.

[5] A. Nierenberg and A. Pasick, “Schools briefing: Coronavirus dorms and super spreaders.” https://www.nytimes.com/2020/09/09/us/schools-reopening-coronavirus.html, 2020. Accessed: 2021-06-01.

[6] D. Ivory, R. Gebeloff, and S. Mervosh, “Young people have less Covid-19 risk, but in college towns, deaths rose fast.” https://www.nytimes.com/2020/12/12/us/covid-colleges-nursing-homes.html, 2021. Accessed: 2021-06-01.

[7] J. Moody, “Colleges extend remote instruction.” https://www.insidehighered.com/news/2022/01/10/colleges-extend-remote-instruction-period-due-omicron, 2022. Accessed: 2022-02-08.

[8] L. Ellis, “Colleges hoped for an in-person fall. Now the dream is crumbling.,” Chronicle of Higher Education, July 20, 2020.

[9] A. D. Paltiel, A. Zheng, and R. P. Walensky, “Assessment of SARS-CoV-2 screening strategies to permit the safe reopening of college campuses in the United States,” JAMA Netw. Open, vol. 3, no. 7, pp. e2016818–e2016818, 2020.

[10] M. S. Wrighton and S. J. Lawrence, “Reopening colleges and universities during the COVID-19 pandemic,” Ann. Intern. Med., 2020. Forthcoming.

[11] R. Callimachi, “For colleges, vaccine mandates often depend on which party is in power.” https://www.nytimes.com/2021/05/22/us/college-vaccine-universities.html, 2021. Accessed: 2021-06-01.

[12] J. V. Lazarus, S. C. Ratzan, A. Palayew, L. O. Gostin, H. J. Larson, K. Rabin, S. Kimball, and A. El-Mohandes, “A global survey of potential acceptance of a COVID-19 vaccine,” Nat. Med., vol. 27, no. 2, pp. 225–228, 2021.

[13] M. Chinazzi, J. T. Davis, M. Ajelli, C. Gioannini, M. Litvinova, S. Merler, A. P. Y Piontti, K. Mu, L. Rossi, K. Sun, et al., “The effect of travel restrictions on the spread of the 2019 novel coronavirus (COVID-19) outbreak,” Science, vol. 368, no. 6489, pp. 395–400, 2020.

[14] M. Gatto, E. Bertuzzo, L. Mari, S. Miccoli, L. Carraro, R. Casagrandi, and A. Rinaldo, “Spread and dynamics of the COVID-19 epidemic in Italy: Effects of emergency containment measures,” Proc. Natl. Acad. Sci. U.S.A, vol. 117, no. 19, pp. 10484–10491, 2020.

[15] M. Gilbert, G. Pullano, F. Pinotti, E. Valdano, C. Poletto, P.-Y. Böelle, E. d’Ortenzio, Y. Yazdanpanah, S. P. Eholie, M. Altmann, et al., “Preparedness and vulnerability of African countries against importations of COVID-19: a modelling study,” Lancet, vol. 395, no. 10227, pp. 871–877, 2020.

[16] H. W. Hethcote, “The mathematics of infectious diseases,” SIAM Review, vol. 42, no. 4, pp. 599–653, 2000.

[17] A. J. Kucharski, T. W. Russell, C. Diamond, Y. Liu, J. Edmunds, S. Funk, R. M. Eggo, and Centre for Mathematical Modelling of Infectious Diseases COVID-19 working group, “Early dynamics of transmission and control of COVID-19: A mathematical modelling study,” Lancet Infect. Dis., vol. 20, pp. 553–558, May 2020.

[18] E. Lofgren, K. Lum, A. Horowitz, B. Madubuonwu, K. Myers, and N. H. Fefferman, “The epidemiological implications of jails for community, corrections officer, and incarcerated population risks from COVID-19,” medRxiv, 2021.

[19] K. Mizumoto and G. Chowell, “Transmission potential of the novel coronavirus (COVID-19) onboard the diamond Princess Cruises Ship, 2020,” Infect. Dis. Model., vol. 5, pp. 264–270, 2020.

[20] P. T. Gressman and J. R. Peck, “Simulating COVID-19 in a university environment,” Math Biosci, vol. 328, p. 108436, 2020.

[21] B. Lopman, C. Liu, A. Le Guillou, A. Handel, T. L. Lash, A. Isakov, and S. Jenness, “A model of COVID-19 transmission and control on university campuses,” medRxiv, 2020.

[22] K. A. Weeden and B. Cornwell, “The small-world network of college classes: implications for epidemic spread on a university campus,” Sociol. Sci., vol. 7, pp. 222–241, 2020.

[23] G. Chowell and F. Brauer, “The basic reproduction number of infectious diseases: computation and estimation using compartmental epidemic models,” in Mathematical and statistical estimation approaches in epidemiology, pp. 1–30, Springer, 2009.

[24] S. B. Patel and P. Patel, “Doubling time and its interpretation for COVID 19 cases,” Natl. J. Community Med, vol. 11, pp. 141–143, 2020.

[25] R. Nunes-Vaz, “Visualising the doubling time of COVID-19 allows comparison of the success of containment measures,” Global Biosecurity, vol. 1, no. 3, 2020.

[26] K. Muniz-Rodriguez, G. Chowell, C.-H. Cheung, D. Jia, P.-Y. Lai, Y. Lee, M. Liu, S. K. Ofori, K. M. Roosa, L. Simonsen, et al., “Doubling time of the COVID-19 epidemic by province, China,” Emerg. Infect. Dis., vol. 26, no. 8, p. 1912, 2020.

[27] A. Saltelli, P. Annoni, I. Azzini, F. Campolongo, M. Ratto, and S. Tarantola, “Variance based sensitivity analysis of model output. design and estimator for the total sensitivity index,” Comput. Phys. Commun., vol. 181, no. 2, pp. 259–270, 2010.

[28] A. Saltelli, M. Ratto, T. Andres, F. Campolongo, J. Cariboni, D. Gatelli, M. Saisana, and S. Tarantola, Global Sensitivity Analysis: The Primer. John Wiley & Sons, 2008.

[29] G. Archer, A. Saltelli, and I. Sobol, “Sensitivity measures, ANOVA-like techniques and the use of bootstrap,” J. Stat. Comput. Simul, vol. 58, no. 2, pp. 99–120, 1997.

[30] T. Homma and A. Saltelli, “Importance measures in global sensitivity analysis of nonlinear models,” Reliab. Eng. Syst. Saf., vol. 52, no. 1, pp. 1–17, 1996.

[31] M. J. Jansen, “Analysis of variance designs for model output,” Comput. Phys. Commun., vol. 117, no. 1-2, pp. 35–43, 1999.

[32] MathWorks, “Statistics and Machine Learning Toolbox<sup>TM</sup> R2020a,” 2020. https://www.mathworks.com/help/stats/.

[33] A. Brlek, Š. Vidovič, S. Vuzem, K. Turk, and Z. Simonović, “Possible indirect transmission of COVID-19 at a squash court, Slovenia, March 2020: case report,” Epidemiol. Infect., vol. 148, 2020.

[34] L. Hamner, “High SARS-CoV-2 attack rate following exposure at a choir practice—Skagit County, Washington, March 2020,” Morbidity and Mortality Weekly Report, vol. 69, 2020.

[35] K.-S. Kwon, J.-I. Park, Y. J. Park, D.-M. Jung, K.-W. Ryu, and J.-H. Lee, “Evidence of long-distance droplet transmission of SARS-CoV-2 by direct air flow in a restaurant in Korea,” J. Korean Med. Sci., vol. 35, no. 46, 2020.

[36] L. M. Glass and R. J. Glass, “Social contact networks for the spread of pandemic influenza in children and teenagers,” BMC Public Health, vol. 8, no. 1, pp. 1–15, 2008.

[37] K. Leung, M. Jit, E. H. Lau, and J. T. Wu, “Social contact patterns relevant to the spread of respiratory infectious diseases in Hong Kong,” Sci. Rep., vol. 7, no. 1, pp. 1–12, 2017.

[38] R. Bahl, N. Eikmeier, A. Fraser, M. Junge, F. Keesing, K. Nakahata, and L. Reeves, “Modeling COVID-19 spread in small colleges,” PLoS One, vol. 16, no. 8, p. e0255654, 2021.

[39] J. Panovska-Griffiths, R. Stuart, C. Kerr, K. Rosenfield, D. Mistry, W. Waites, D. Klein, C. Bonell, and R. Viner, “Modelling the impact of reopening schools in the UK in early 2021 in the presence of the alpha variant and with roll-out of vaccination against SARS-CoV-2,” J. Math. Anal. Appl., p. 126050, 2022.

[40] Y. Liu, A. A. Gayle, A. Wilder-Smith, and J. Rocklöv, “The reproductive number of COVID-19 is higher compared to SARS coronavirus,” J. Travel Med., 2020.

[41] M. Park, A. R. Cook, J. T. Lim, Y. Sun, and B. L. Dickens, “A systematic review of COVID-19 epidemiology based on current evidence,” J. Clin. Med., vol. 9, no. 4, p. 967, 2020.

[42] R. Ke, E. Romero-Severson, S. Sanche, and N. Hengartner, “Estimating the reproductive number R0 of SARS-CoV-2 in the United States and eight European countries and implications for vaccination,” J Theor Biol, vol. 517, p. 110621, 2021.

[43] S. A. Lauer, K. H. Grantz, Q. Bi, F. K. Jones, Q. Zheng, H. R. Meredith, A. S. Azman, N. G. Reich, and J. Lessler, “The incubation period of coronavirus disease 2019 (COVID-19) from publicly reported confirmed cases: estimation and application,” Ann. Intern. Med., vol. 172, no. 9, pp. 577–582, 2020.

[44] J. A. Backer, D. Klinkenberg, and J. Wallinga, “Incubation period of 2019 novel coronavirus (2019-nCoV) infections among travellers from Wuhan, China, 20–28 January 2020,” Eurosurveillance, vol. 25, no. 5, p. 2000062, 2020.

[45] A. McCombs and C. Kadelka, “A model-based evaluation of the efficacy of COVID-19 social distancing, testing and hospital triage policies,” PLoS Comput. Biol., vol. 16, no. 10, p. e1008388, 2020.

[46] M. Casey-Bryars, J. Griffin, C. McAloon, A. Byrne, J. Madden, D. Mc Evoy, Á. Collins, K. Hunt, A. Barber, F. Butler, et al., “Presymptomatic transmission of SARS-CoV-2 infection: a secondary analysis using published data,” BMJ Open, vol. 11, no. 6, p. e041240, 2021.

[47] D. P. Oran and E. J. Topol, “The proportion of SARS-CoV-2 infections that are asymptomatic: a systematic review,” Ann. Intern. Med., vol. 174, no. 5, pp. 655–662, 2021.

[48] P. Sah, M. C. Fitzpatrick, C. F. Zimmer, E. Abdollahi, L. Juden-Kelly, S. M. Moghadas, B. H. Singer, and A. P. Galvani, “Asymptomatic SARS-CoV-2 infection: A systematic review and meta-analysis,” Proc. Natl. Acad. Sci. U.S.A, vol. 118, no. 34, p. e2109229118, 2021.

[49] D. Buitrago-Garcia, A. M. Ipekci, L. Heron, H. Imeri, L. Araujo-Chaveron, I. Arevalo-Rodriguez, A. Ciapponi, M. Cevik, A. Hauser, M. I. Alam, et al., “Occurrence and transmission potential of asymptomatic and presymptomatic SARS-CoV-2 infections: Update of a living systematic review and meta-analysis,” PLoS Medicine, vol. 19, no. 5, p. e1003987, 2022.

[50] R. Li, S. Pei, B. Chen, Y. Song, T. Zhang, W. Yang, and J. Shaman, “Substantial undocumented infection facilitates the rapid dissemination of novel coronavirus (SARS-CoV-2),” Science, vol. 368, no. 6490, pp. 489–493, 2020.

[51] J. P. Townsend, H. B. Hassler, P. Sah, A. P. Galvani, and A. Dornburg, “The durability of natural infection and vaccine-induced immunity against future infection by SARS-CoV-2,” Proc. Natl. Acad. Sci. U.S.A, vol. 119, no. 31, p. e2204336119, 2022.

[52] J. Pan, C. Harb, W. Leng, and L. C. Marr, “Inward and outward effectiveness of cloth masks, a surgical mask, and a face shield,” Aerosol Sci. Technol., vol. 55, no. 6, pp. 718–733, 2021.

[53] J. Howard, A. Huang, Z. Li, Z. Tufekci, V. Zdimal, H.-M. van der Westhuizen, A. von Delft, A. Price, L. Fridman, L.-H. Tang, V. Tang, G. L. Watson, C. E. Bax, R. Shaikh, F. Questier, D. Hernandez, L. F. Chu, C. M. Ramirez, and A. W. Rimoin, “An evidence review of face masks against COVID-19,” Proc. Natl. Acad. Sci. U.S.A, vol. 118, no. 4, 2021.

[54] Q. Bi, Y. Wu, S. Mei, C. Ye, X. Zou, Z. Zhang, X. Liu, L. Wei, S. A. Truelove, T. Zhang, et al., “Epidemiology and transmission of COVID-19 in 391 cases and 1286 of their close contacts in Shenzhen, China: a retrospective cohort study,” Lancet Infect. Dis., vol. 20, no. 8, pp. 911–919, 2020.

[55] R. Verity, L. C. Okell, I. Dorigatti, P. Winskill, C. Whittaker, N. Imai, G. Cuomo-Dannenburg, H. Thompson, P. G. Walker, H. Fu, et al., “Estimates of the severity of coronavirus disease 2019: a model-based analysis,” Lancet Infect. Dis., vol. 20, no. 6, pp. 669–677, 2020.

[56] F. Zhou, T. Yu, R. Du, G. Fan, Y. Liu, Z. Liu, J. Xiang, Y. Wang, B. Song, X. Gu, et al., “Clinical course and risk factors for mortality of adult inpatients with COVID-19 in Wuhan, China: a retrospective cohort study,” Lancet, vol. 395, no. 10229, pp. 1054–1062, 2020.

[57] M. W. Tenforde, S. S. Kim, C. J. Lindsell, E. Billig Rose, N. I. Shapiro, D. C. Files, K. W. Gibbs, H. L. Erickson, J. S. Steingrub, H. A. Smithline, et al., “Symptom duration and risk factors for delayed return to usual health among outpatients with COVID-19 in a multistate health care systems network—United States, March–June 2020,” Morbidity and Mortality Weekly Report, vol. 69, no. 30, pp. 993–998, 2020.

[58] Y.-H. Lee, C. M. Hong, D. H. Kim, T. H. Lee, and J. Lee, “Clinical course of asymptomatic and mildly symptomatic patients with coronavirus disease admitted to community treatment centers, South Korea,” Emerg. Infect. Dis., vol. 26, no. 10, p. 2346, 2020.

